# Diversity and within-host evolution of parasites from VL and VL/HIV patients in Northern Ethiopia

**DOI:** 10.1101/2021.04.01.21254750

**Authors:** Susanne U. Franssen, Yegnasew Takele, Emebet Adem, Mandy J. Sanders, Ingrid Müller, Pascale Kropf, James A. Cotton

## Abstract

Visceral leishmaniasis (VL) is a fatal disease and a growing public health problem in East Africa, where Ethiopia has one of the highest VL burdens. The largest focus of VL in Ethiopia is driven by high prevalence in migrant agricultural workers and associated with a high rate of co-infection with HIV. This co-infection makes VL more difficult to treat successfully, and is associated with a high rate of relapse, with VL/HIV patients frequently experiencing many relapses of VL before succumbing to this infection. We present genome-wide data on *Leishmania donovani* isolates from a longitudinal study of cohorts of VL and VL/HIV patients reporting to a single clinic in Ethiopia. Extensive clinical data allows us to investigate the influence of co-infection and relapse on the populations of parasites infecting these patients. We find that the same parasite population is responsible for both VL and VL/HIV infections, and that in most cases, disease relapse is caused by recrudescence of the population of parasites that caused primary VL. Complex, multi-clonal infections are present in both primary and relapse cases, but the infrapopulation of parasites within a patient loses genetic diversity between primary disease presentation and subsequent relapses, presumably due to a population bottleneck induced by treatment. These data suggest that VL/HIV relapses are not caused by genetically distinct parasite infections, nor by re-infection. Treatment of VL does not lead to sterile cure, and in VL/HIV the infecting parasites are able to re-establish after clinically successful treatment, leading to repeated relapse of VL.

**Importance:** Visceral leishmaniasis (VL) is the second largest cause of deaths due to parasite infections, and a growing problem in East Africa. In Ethiopia, it is particularly associated with migrant workers moving from non-endemic regions for seasonal agricultural work, and frequently found as a co-infection with HIV, which leads to frequent VL relapse following treatment. Insight into the process of relapsing in these patients is thus key to controlling the VL epidemic in Ethiopia. We show that there is little genetic differentiation between the parasites infecting HIV positive and HIV negative VL patients. Moreover, we provide evidence that relapses are caused by the initially infecting parasite population, and that treatment induces a loss of genetic diversity in this population. We propose that restoring functioning immunity and improving anti-parasitic treatment may be key in breaking the cycle of relapsing VL in VL/HIV patients.

## Introduction

Leishmaniasis is a major neglected tropical disease, with the most severe form – visceral leishmaniasis (VL) – being the second most deadly parasitic disease, responsible for 20,000 to 40,000 deaths per year (Alvar et al. 2012), together with significant morbidity (loss of over 2 million DALYs – disability-adjusted life years; Mathers, Ezzati, and Lopez 2007). VL is caused by infections with protozoan parasites from the *Leishmania donovani* species complex. While clinical VL is frequently fatal without adequate treatment, most *L. donovani* infections remain asymptomatic (see review in Chappuis et al. 2007; Singh et al. 2014), or at least subclinical, and much remains unknown about what leads to the development of severe disease (McCall, Zhang, and Matlashewski 2013) or to the success or failure of treatment. Most of the global burden of VL is in the Indian subcontinent, but East Africa represents the second largest burden of VL caused by *L. donovani*. While the number of cases has been falling in the Indian subcontinent (Rijal et al. 2019), they continue to increase in Africa. Ethiopia, Sudan and South Sudan have the highest prevalence in this region, and VL is one of the most significant vector-borne diseases in Ethiopia, with over 3 million people at risk of infection in a number of distinct foci in lowland areas of the country (Gadisa et al. 2015; Leta et al. 2014).

The most important focus of VL in Ethiopia is around Metema and Humera in the Northwest of the country, where since the 1970s there has been an important focus of disease particularly associated with seasonal migration of non-immune labourers from the surrounding non-endemic highland regions for agricultural work. Several outbreaks have occurred in this focus, and there is evidence that this focus is spreading and has seeded outbreaks in other, previously non-endemic areas (Gadisa et al. 2015; Leta et al. 2014; Gelanew et al. 2011; Alvar et al. 2007). One factor in the growth of VL in Ethiopia is co-infection, and particularly co-infection of *L. donovani* with HIV. While VL/HIV coinfection is a widespread concern (Lindoso et al. 2018), Ethiopia has the highest rate of VL/HIV co-infections in Africa – and possibly globally – and while estimates vary between studies, HIV may be present in over 20% of VL cases (Leta et al. 2014; Mohebali and Yimam 2020). In Ethiopia and elsewhere, HIV coinfection is known to increase the probability that *L. donovani* infections progress to symptomatic visceral leishmaniasis (Hurissa et al. 2010; Albuquerque et al. 2014). These co-infections are difficult to treat, with poor clinical outcomes for patients with very high mortality and high rates of relapse following treatment (Hurissa et al. 2010; Diro et al. 2019; Alemayehu, Wubshet, and Mesfin 2016; Diro et al. 2014; Mohammed et al. 2020). It is generally thought that cure of VL is not sterile, so that parasites continue to be present, but there is little direct evidence of this. It is unknown whether relapses of VL are due to re-infection by parasites – which we would expect to occur more commonly when patient immunity is compromised – or due to recrudescence of the original infection from parasites that persist through original treatment.

There has been significant work on the genetics of *L. donovani* in East Africa, which has revealed Ethiopia as a hotspot for diversity of visceral leishmaniasis, with two major genetic groups of *L. donovani* occurring (Gelanew et al. 2010). These groups were shown to be transmitted by distinct sandfly vectors and are separated by the rift valley (Teshome Gebre-Michael et al. 2010; T. Gebre-Michael and Lane 1996). A single Ethiopian isolate has been identified from a third group of parasites, related to those found on the Arabian Peninsula and adjacent areas of the Middle East (Franssen et al. 2020). To add to this complex picture, a 2004 outbreak of VL in northern Ethiopia has been associated with a diverse population of parasites that persisted after the outbreak (Gelanew et al. 2011) some of which are hybrids between two of these groups (Cotton et al. 2020; Gelanew et al. 2014, 2010). However, the studies to-date represent relatively small numbers of isolates often collected over an extended period of time and from a wide geographical area. While this has established an important baseline in understanding the diversity and basic structure of parasite populations, these studies give little or no insight into the origins of the observed genetic variation or its phenotypic consequences. For example, any genetic variation driving variation in disease presentation or clinical outcome is confounded with variation over time and space.

Here, we report whole-genome sequence data from parasites isolated from two cohorts, VL patients and patients coinfected with HIV and *Leishmania*. All patients were recruited at a single clinic in Northern Ethiopia over a period of 18 months. This makes our work one of the largest studies of genome-wide genetic variation in leishmaniasis at a single place and time. Extensive clinical and immunological data was collected from these patients over 12 months of follow-up, and long-term outcomes recorded up to 3 years post VL diagnosis, so the parasite genome data we report is from a well-understood patient population. Full details of patient recruitment and clinical data from these patients have previously been reported (Takele et al. 2021). Parasite isolation is generally only possible when either bone marrow or splenic aspirates are being performed for parasitological investigation, which is routinely done at diagnosis of each VL episode. This includes additional isolates from individual patients that were collected where possible during subsequent episodes of clinical relapse during the follow-up of this cohort. We thus present – to our knowledge – the first longitudinal data on within-host evolution of *Leishmania* parasites. These data demonstrate that most relapse cases are caused by the originally infecting parasites, and that the parasite infrapopulations (the population of parasites within a host at a particular time) undergo strong bottlenecks during treatment. VL relapse is not generally associated with known markers for drug resistance, and despite some diversity, we show that parasites from VL infections without HIV and VL/HIV co-infections represent the same parasite population. We conclude that relapse in VL/HIV patients is due to the persistence of parasites after treatment, that in the absence of fully functional host immunity can re-expand to cause the recurrent episodes of disease.

## Results

### No distinct phylogenetic origin of parasites from VL and VL/HIV patients in Northern Ethiopia

We sequenced the genomes of 108 *L. donovani* isolates from a total of 98 VL patients in Ethiopia. For five isolates, two aliquots of the same initial culture were processed and sequenced independently. The genome-wide haploid coverage of the sequenced samples ranged between 7 and 30 (median 13). Our study included 68 patients, who had VL only, and 29 patients were additionally coinfected with HIV and were receiving antiretroviral treatment (Takele et al. 2021). The complete metadata of samples taken, technical replicates and patients is summarised in table S1.

These parasites were isolated from a cohort of VL patients in which 78.1% of the successfully treated VL/HIV patients relapsed within a three-year time period, while none of the patients without HIV infection relapsed (Takele et al. 2021). Therefore, we tested for the possibility that there is a genetic difference between parasites isolated from HIV negative vs. HIV positive VL patients that could explain this difference in relapse rate. Phylogenetic reconstruction using the first parasite isolate taken from each patient – which is not necessarily the first episode of VL in the HIV patients – shows the genetic relatedness between isolates based on whole-genome SNP data (Fig. 1A). It suggested that there was no clear phylogenetic structure between parasites from the two groups. To investigate more subtle genetic structure, we used two measures to estimate a possible association of HIV status with genome-wide parasite genomics, based on those isolates from 67 HIV negative and 29 HIV positive patients, respectively. Analysis of similarities (ANOSIM) using pairwise genome-wide Nei’s D as distance measure (R=0.09321, p-value=0.0290, FDR=0.048) as well as Blomberg’s K (K=0.0348, p-value=0.048, FDR=0.048), both showed an association of HIV status with the phylogeny at α=0.05 but not at α=0.01. Isolates from HIV positive patients were more frequently from second or subsequent relapses of VL disease, which could introduce a bias due to a longer period of within-host evolution in these infections. Therefore, we also tested for the association of HIV status based on isolates taken during the first episode of VL disease for a particular patient (Figure S1; 67 samples from HIV negative and 5 samples from HIV positive patients). In this case no significant association was present (ANOSIM: R=-0.0602, p-value=0.6; Blomberg’s K: K=0.0197, p-value=0.4).

**Figure 1.**
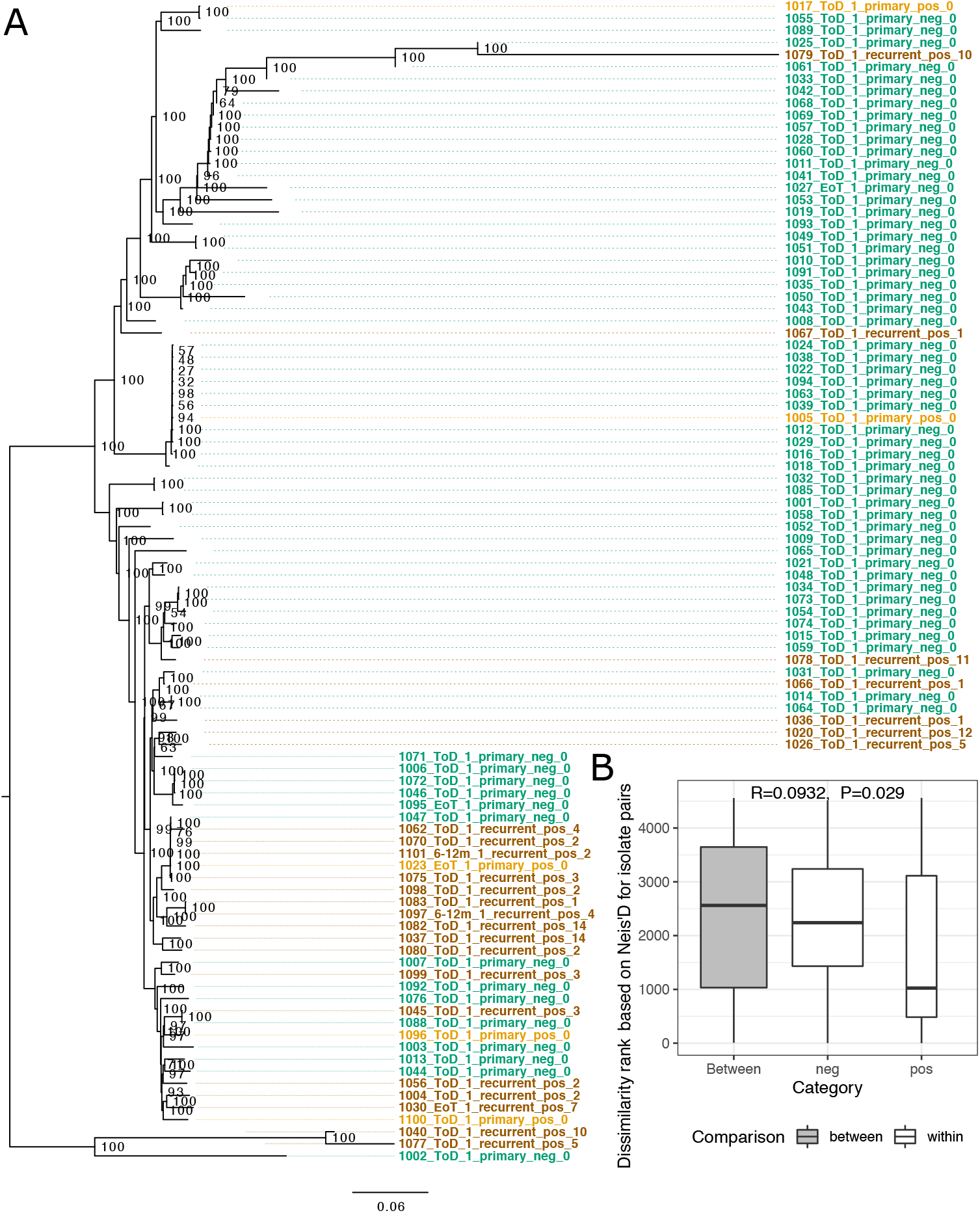
Genomic relatedness between first parasite isolates taken from each patient. A) Phylogeny of first isolate taken from each patient. Parasite sample names are encoded including the patient number, treatment status, patient isolate count, VL type, HIV status and the VL episode number of the patient the isolate was taken. Sample colour indicates VL and HIV status: green for primary VL episodes in HIV negative patients, light orange for primary VL episodes in HIV positive patients and ochre for samples from VL relapse in VL/HIV coinfection cases. Bootstrap values are indicated at branch nodes. The phylogeny is rooted based on the inclusion of an *L. infantum* outgroup (data not shown). B) ANOSIM results comparing the sum of ranked pairwise genetic distances (Nei’s D) within and between HIV positive and negative samples shown in the phylogeny.

### Recurrent VL in HIV patients is caused by persistent parasites

Seven of the patients in our study had multiple parasite isolates taken as part of clinical investigation of VL relapses during the first 12 months follow-up period (Table 1). For the remaining 92 patients, parasite isolates could only be taken at a single time point during our study either due to the absence of relapses, or because taking additional bone marrow or splenic aspirates was not clinically justified. To identify if recurrent VL had been caused by re-expansion of persistent parasites within a patient after initial cure, we compared Nei’s genetic distances based on whole genome SNP data between all pairwise sample combinations. Longitudinal samples of parasites from a patient are typically most closely related to parasites samples from the same patient at different times (Fig. 2A). For two time series samples, (1023_EoT_1_primary_pos_0 and 1045_ToD_1_recurrent_pos_3) the genetically most similar samples came from another patient (Fig. 2A, S2A, B). For patient 1023, the genetic distance between samples within the patient gradually increased with time, while the first sample of this patient was very closely related to isolates from two other patients. However, for patient 1045 parasites isolated from the third VL episode were very different from parasites isolated seven months later during the fourth recurrent VL episode, being more closely related to several isolates from other patients. This trend was confirmed when comparing sequential isolates from a patient to the initial isolate from the same patient (Fig. 2B). While within-host parasite changes gradually increased with time, those genetic distances were still very small compared to the median pairwise genetic distance in our entire data set. In this context, two isolates taken just before and after treatment of the fourth VL relapse for patient 1045, were clearly outliers, with a genetic distance similar to pairs of samples from different patients, indicating possible reinfection only in this case (Fig. 2B). Together this suggests that the majority (6 out of 7) of HIV coinfected VL patients relapsed due to the original persistent parasite strain rather than reinfection with *L. donovani*.

**Table 1.**
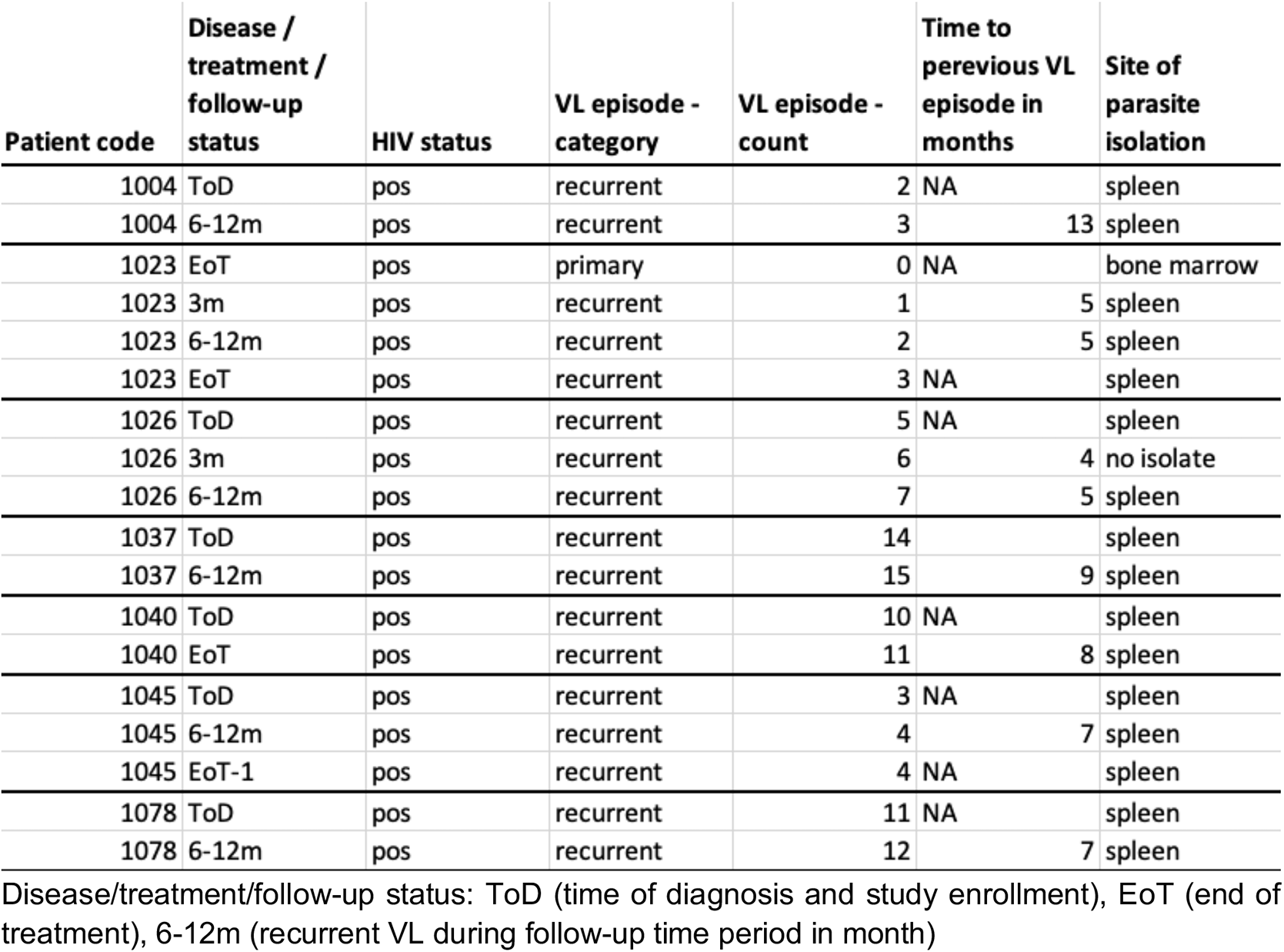
Summary of isolates from patients with multiple samples over time.

| Patient code | Disease /<br>treatment /<br>follow-up<br>status | HIV status | VL episode -<br>category | VL episode -<br>count | Time to<br>previous VL<br>episode in<br>months | Site of<br>parasite<br>isolation |
| --- | --- | --- | --- | --- | --- | --- |
| 1004 | ToD | pos | recurrent | 2 | NA | spleen |
| 1004 | 6-12m | pos | recurrent | 3 | 13 | spleen |
| 1023 | EoT | pos | primary | 0 | NA | bone marrow |
| 1023 | 3m | pos | recurrent | 1 | 5 | spleen |
| 1023 | 6-12m | pos | recurrent | 2 | 5 | spleen |
| 1023 | EoT | pos | recurrent | 3 | NA | spleen |
| 1026 | ToD | pos | recurrent | 5 | NA | spleen |
| 1026 | 3m | pos | recurrent | 6 | 4 | no isolate |
| 1026 | 6-12m | pos | recurrent | 7 | 5 | spleen |
| 1037 | ToD | pos | recurrent | 14 |  | spleen |
| 1037 | 6-12m | pos | recurrent | 15 | 9 | spleen |
| 1040 | ToD | pos | recurrent | 10 | NA | spleen |
| 1040 | EoT | pos | recurrent | 11 | 8 | spleen |
| 1045 | ToD | pos | recurrent | 3 | NA | spleen |
| 1045 | 6-12m | pos | recurrent | 4 | 7 | spleen |
| 1045 | EoT-1 | pos | recurrent | 4 | NA | spleen |
| 1078 | ToD | pos | recurrent | 11 | NA | spleen |
| 1078 | 6-12m | pos | recurrent | 12 | 7 | spleen |

Disease/treatment/follow-up status: ToD (time of diagnosis and study enrollment), EoT (end of treatment), 6-12m (recurrent VL during follow-up time period in month)

**Figure 2.**
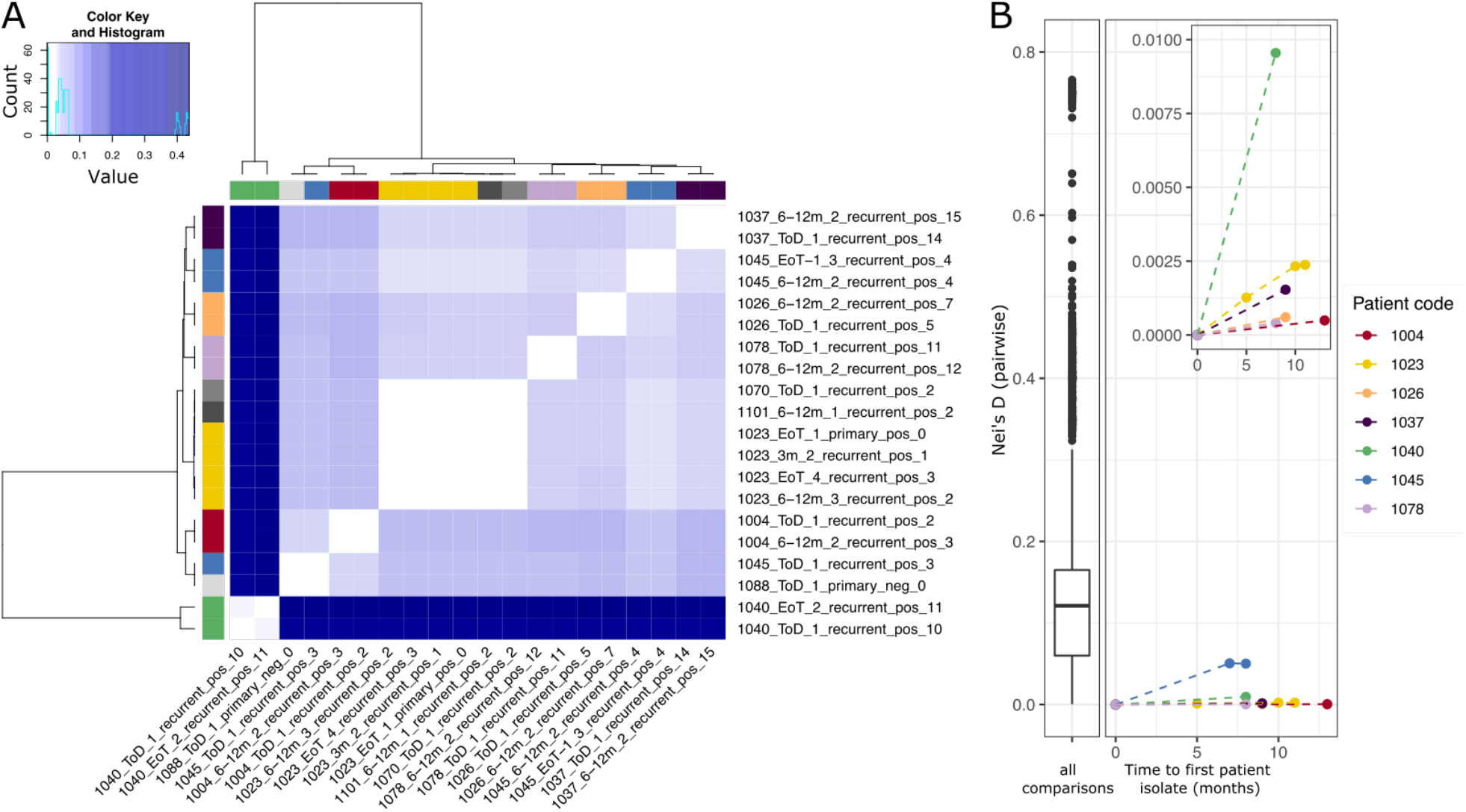
Recurrent VL is predominantly caused by persistent parasites not reinfection. For seven patients parasite isolates were taken at multiple time points during the course of their VL infection (Table 1). A) Heatmap of pairwise Nei’s distances between isolates from the seven patients with longitudinal sampling along with any other isolates from our cohort most closely related to those. Row- and column-side colours indicate patient origin, with colours as used in panel B, except that grey indicates isolates from patients represented by only a single isolate. B) Increase of genetic distance between the first parasite isolate from a patient and subsequent isolates. The left panel shows the distribution of pairwise genetic distances across all 106 parasite isolates, excluding technical replicates and isolates with incomplete metadata. The right panel shows the pairwise Nei’s distances between the first isolate for each patient with longitudinal samples and subsequent samples from that patient (y axis), plotted against the time in months between sampling dates (x axis). Samples are coloured by patient origin.

### Reduction in parasite heterozygosity during within-host evolution

As persistent *Leishmania* populations seem to play a dominant role in recurrent VL in HIV positive patients, we investigated whether particular patterns of genomic change might be associated with this prolonged within-host evolution. Ideally, we would compare longitudinal samples within the same patient over multiple relapses of VL, but the ethical constraints on taking appropriate isolates imply that relatively few such sample sets could be taken (see table 1). The largest fraction of our parasite samples were each isolated from different patients with primary VL and at different numbers of recurrent VL episodes. The identification of convergent genetic changes that could indicate functionally relevant adaptations to prolonged immune challenge is more challenging from these cross-sectional data as changes will arise in different genomic backgrounds and within-host adaptation will be conflated with genetic variation that was present in the initially infecting strain.

We thus first inspected more general patterns of genomic change with recurrent VL. The isolates represent samples of the diversity of parasites present in the infected tissue. From a previous analysis it is clear that diploid SNP variants called by GATK can show allele frequencies far from the 0.5 expected for a diploid genotype (Franssen et al. 2020). We therefore reasoned that the proportion of variable sites called as heterozygotes will be a good approximation of the genetic diversity of the infecting infrapopulation and give some indication of the effective population size. While we refer to this measure as ‘heterozygosity’ it represents a combination of heterozygous differences within individual parasite clones and genetic differences within a diverse and potentially polyclonal infection. In our data, heterozygosity was much lower in isolates from recurrent VL in HIV positive patients than in primary VL in HIV negative patients (Fig. 3A, ANOVA F=16.25, p-value=8.52*10^-7; TukeyHSD for recurrent_pos vs. primary_neg, FDR=0.0000004; recurrent_pos vs. primary_pos, FDR=0.0893283; primary_pos vs. primary_neg, FDR=0.8705747). Isolates from primary episodes of VL in HIV infected patients had intermediate levels of heterozygosity, with a greater difference of the “primary_pos” cohort to the “recurrent_pos” than to the “primary_neg” cohort. This analysis left it unclear whether reduction in heterozygosity was associated with multiple episodes of VL disease only or if differences between the primary VL in HIV positive and HIV negative patients were also present. Therefore, we tested whether the fraction of heterozygous sites in primary VL isolates varied with HIV infection status when the reported length of illness before parasite isolation or the estimated parasite load at the time of isolation were taken into account. However, neither of these covariates had a significant effect on heterozygosity, and no difference between HIV categories was observed (Fig. S3A,B).

**Figure 3.**
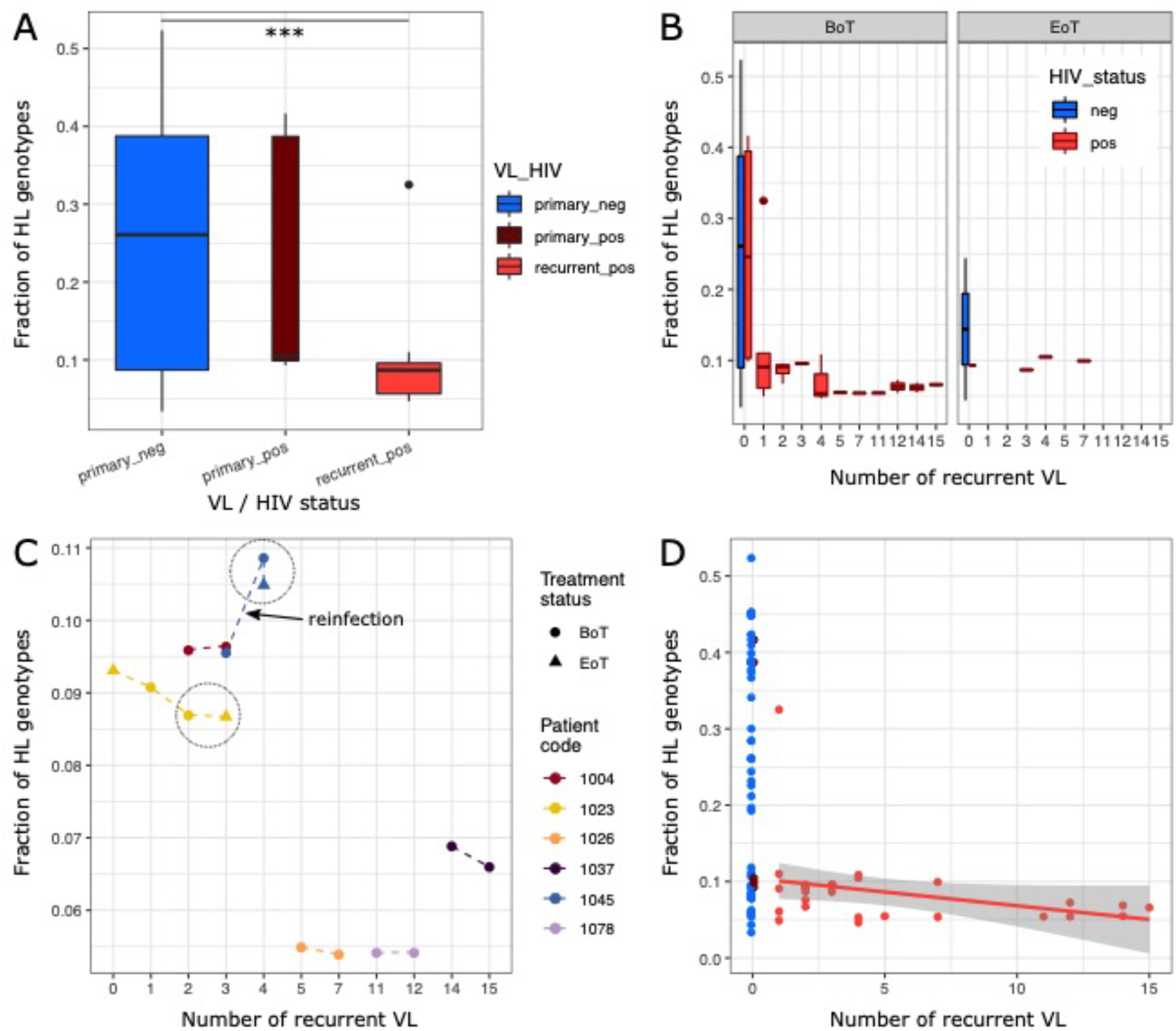
Parasite genomic changes with recurrent VL. A) Heterozygosity (proportion of variable loci called as heterozygous genotypes; HL) in isolates from primary VL in HIV negative patients (primary_neg), primary VL in HIV infected patients (primary_pos) and relapse VL episodes in HIV positive patients (recurrent_pos). There is a significant difference in heterozygosity between “primary_neg” and “recurrent_pos” (ANOVA, TukeyHSD ***: FDR<0.001). B) Heterozygosity values (as shown in A) shown for each number of episodes of VL relapse for isolates taken before (BoT) and at the end (EoT) of treatment for that VL episode. C) Fraction of heterozygous (HL) genotypes for samples from patients with multiple isolates taken. The large black circles group isolates from consecutive sampling events for a patient before and after treatment (dots indicate samples isolated before (BoT) and triangles at the end of treatment (EoT)). The line segment connecting the two isolates suspected to be a reinfection event is indicated (see Fig. 2). D) The fraction of heterozygous genotypes categorised by VL and HIV status and shown against the number of recurrent VL across patients. A linear model fit is shown for isolates from the HIV positive, VL relapse cohort.

Next, we investigated the timing of the reduction in heterozygosity over the course of primary and relapse VL episodes. In the cross-sectional comparison, there was a strong difference in heterozygosity between isolates from patients with primary infections and those experiencing the first relapse of VL (Fig. 3A), suggesting that treatment of primary VL itself already reduced heterozygosity levels. To test this directly, we looked at how heterozygosity of parasite isolates varied with the number of previous VL episodes and compared samples taken before and after treatment and in both HIV positive and negative patients (Fig. 3B). For the primary VL data, all three post-treatment samples suggested a noticeable reduction in heterozygosity levels compared to before treatment isolates, but the low sample size impeded assessing statistical significance (Fig. 3B). Moreover, a small treatment effect was also still present for recurrent VL in our time series data (Fig. 3C, patients 1023 and 1045). While the reduction in heterozygosity between primary VL and the first relapse of VL was strongest, there was a trend for a linear decrease in heterozygosity of small effect size from the first VL relapse and subsequent recurrence of VL though only significant at a lenient threshold of α=0.1 (linear model: fHL ∽ num_of_relapses, p-value=0.0798, Fig. 3D). This relationship was also reflected in our within-patient longitudinal data, where parasite heterozygosity levels decreased through time with additional relapse episodes. The only exception was a clear increase for patient 1045, where we had previously suggested reinfection (Fig. 3C). Taken together, these data suggest a strong effect of initial drug treatment on heterozygosity levels, followed by a weaker continuous effect, presumably due to repeated population size decreases during drug treatment.

### Infections in recurrent VL are as complex as primary infections

The observed reduction of heterozygosity with recurrent VL could have been partly due to clonal diversity compared to a heterozygous allele shared across all cells in a population. Therefore, we investigated the amount of possible clonal diversity (the complexity of infection) in our isolates. By inspecting the distributions of minor allele frequencies at diploid chromosomes, we categorised them into isolates likely to come from single clonal infections (Fig. S4A), isolates potentially representing complex infections with multiple clones (Fig. S4C) and those with slightly noisy allele frequency distributions that either represented very few variants or had a few variants present at unexpected frequencies (Fig. S4B, S5). Despite this somewhat subjective assessment, complex infections were clear outliers in terms of mean distance from the expected allele frequency and the variance of observed frequencies (Fig. S4D). Complex infections were found in 9% (6/68) of our primary isolates and 14% (4/29, one additional technical replicate) of the isolates from recurrent VL. None of our within-patient longitudinal data gained complexity with time but one lost complexity between recurrent VL episode 1 vs. 2 (patient 1040). The four complex isolates from recurrent VL are from episodes 1, 2 and twice from episode 10. Noisy profiles were more common in isolates from recurrent VL likely due to the small number of heterozygous sites in these isolates that reduced the signal-to-noise ratio. If the reduced heterozygosity we observe in recurrent VL was due to loss of clones in complex infections, we would expect complex infections to be common in primary isolates, and less common in isolates from recurrent VL, which was not the case for our samples.

### Unclear role of variation in chromosome dosage in within host heterozygosity reduction

*Leishmania* species are known to vary widely in chromosome copy number, with even different cells within a single clonal population varying in chromosome dosage (termed mosaic aneuploidy). This characteristic is generated through rapid change in chromosome copy number between mother and daughter cells for a subset of chromosomes, leading to cells having different aneuploidy profiles of chromosome dosage. Reductions in copy number are expected to reduce heterozygosity of individual parasites (Sterkers et al. 2012). Moreover, frequent turnover of chromosome copies should lead to a reduction in observed strain allelic diversity if a particular haplotype is favoured by selection or drift is high due to small population sizes. While previous work suggests that aneuploidy mosaicism and turnover is rare in the mammalian host compared to promastigote culture (Dumetz et al. 2017), it could still contribute to the observed reduction of heterozygosity during within-host evolution. As we only sequenced DNA extracted from pools of cells for each parasite isolate instead of individual cells, we cannot observe cell-to-cell variation. Therefore, we compared aneuploidy profiles between isolates from our time series data as well as for independently sequenced aliquots from some isolates as a conservative estimate of the degree of aneuploidy variation in our study (Table S2). The aneuploidy level was generally low in our sample cohort with 73.5% (83/113) of all samples having a population aneuploidy profile of diploid chromosomes apart from a tetraploid chromosome 31 (Fig. S6). Time series samples and replicate primary isolates were from a total of 11 patients. 72.7% (8/11) of those patients with multiple samples had the same population-wide aneuploidy profiles in all samples. In contrast, isolates from patients 1023, 1037 and 1045 showed some aneuploidy variation either between sampling events or between aliquots from the same biopsy sample (Fig. 4, Table 2, S2). This suggests that a low level of aneuploidy variation is present within patients though we cannot definitively associate this variation with the reduction in heterozygosity we observe in isolates from patients with repeated relapses.

**Table 2:**
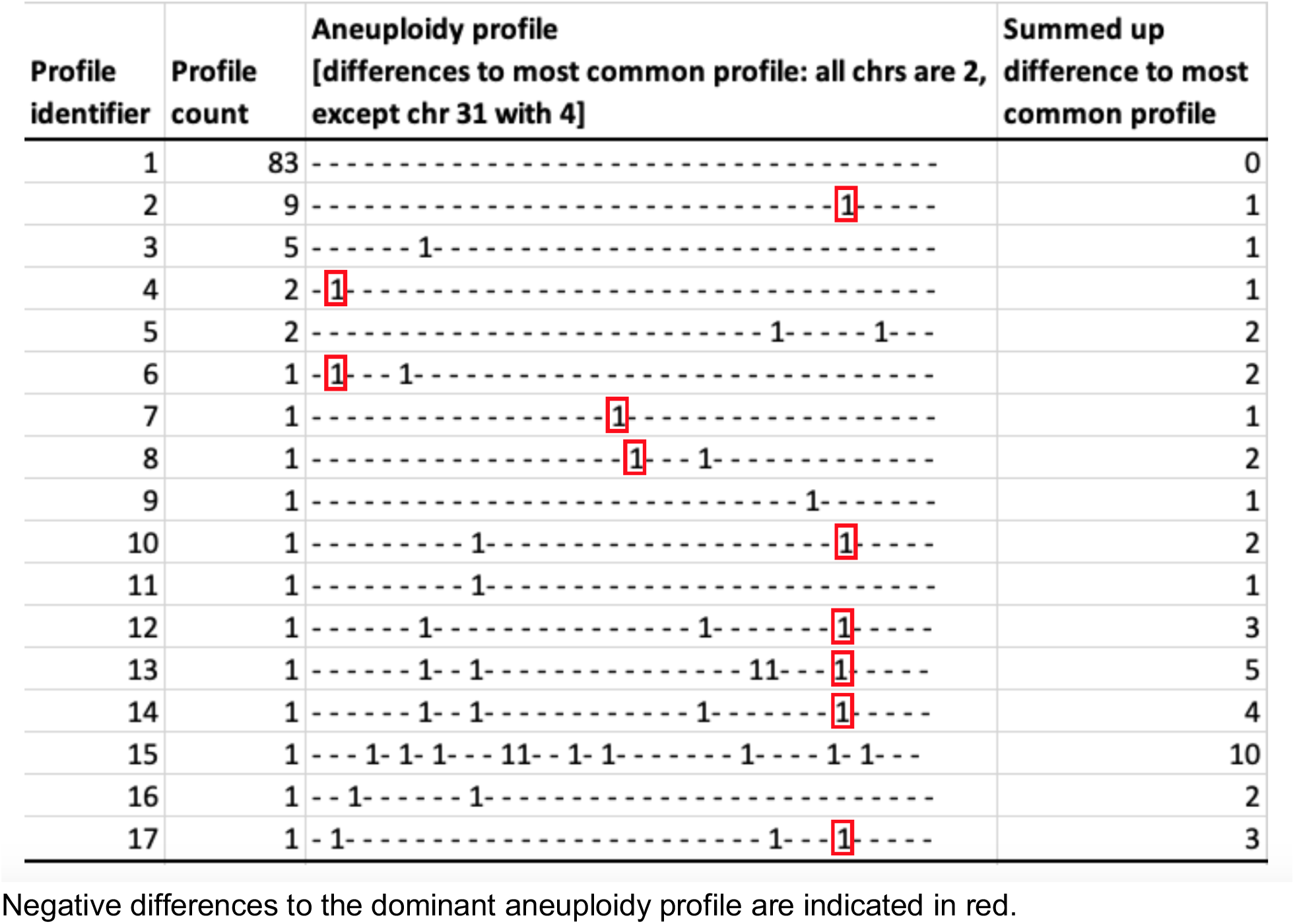
Summary of aneuploidy profiles and their abundances across all parasite isolates.

**Figure 4.**
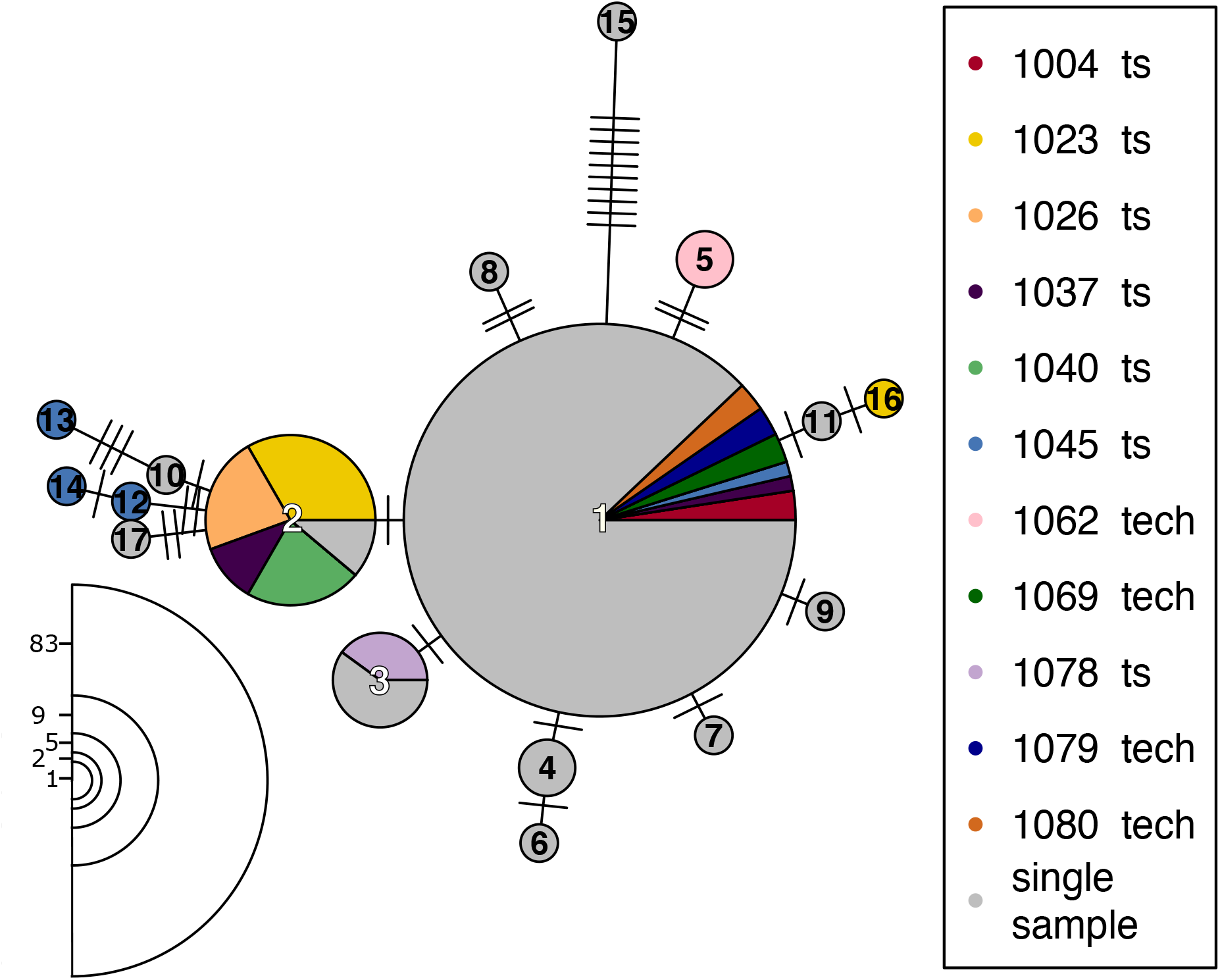
Network of aneuploidy profiles. Aneuploidy profiles of all 113 parasite isolates are shown in a minimum spanning network. Each circle represents a distinct aneuploidy profile and edges between circles indicate similarity between profiles, with the number of edge ticks indicating the total somy difference between profiles. Colours indicate patient origin, with patients represented by only single isolates all gray and those with either multiple temporal samples (ts) or with aliquoted isolates (tech) in unique colours. Circle sizes represent the numbers of isolates showing a particular profile and numbers for each circle size are indicated in the left lower corner. Circles are numbered to identify each aneuploidy profile, and details of the somy patterns and abundance of each profile are listed in table 2. The largest circle (profile 1) represents the diploid condition, but with a tetrasomic chromosome 31.

### No signal of drug resistance evolution during repeated treatment of recurrent VL

A possible explanation for the survival of parasites through treatment is the evolution of drug resistance within the parasite infrapopulation, which would also contribute to the high rate of treatment failure during subsequent VL relapses. However, so far there is little direct evidence that treatment failure in VL/HIV patients is largely driven by drug resistance. We investigated whether known markers for drug resistance in *Leishmania* were present in our study cohorts and compared their frequencies between primary and recurrent VL. Gene copy numbers were estimated for genes within four loci previously suggested to be involved in drug resistance including the H-locus (Callahan and Beverley 1991), the M-locus (MAPK1 gene; Ashutosh et al. 2012), both associated with resistance to antimonial drugs, the Miltefosine transporter (Pérez-Victoria et al. 2003) and the Miltefosine sensitivity locus (MSL; Carnielli et al. 2018). We tested for change in copy number at each of these four loci in HIV negative primary VL cases (“primary_neg”), HIV positive primary cases (“primary_pos”) and from relapse cases in HIV positive patients (“recurrent_pos”). Several genes showed increase with respect to the baseline chromosome copy number for the two larger groups (“primary_neg” and “recurrent_pos”) – including YIP, MRPA and ASS at the H-locus, and at a gene associated with the Miltefosine transporter (annotated as a hypothetical protein in the LV9 reference genome; one sample t-tests, FDR<0.001, Fig. 5); only in two of these above cases the “primary_pos” cohort had a significant increase in copy number compared to the chromosome background, but this cohort also had far fewer samples making it hard to detect small effect size changes (one sample t-tests, FDR<0.05, Fig. 5). Comparison between all three groups for each gene did not show any differences between the three disease categories except for the second MAPK1 ortholog (ANOVA, FDR<0.05, Fig. 5). The increased dosage at the H-locus in our cohort is in line with previous findings from Ethiopian isolates (Franssen et al. 2020), but as both - this increase as well as the dosage increase of the Miltefosine transporter-associated gene - are similarly present in both HIV positive and negative patients, they cannot explain VL relapse. Only for the second ortholog of the MAPK1 gene does our data support a copy number increase with relapse compared to primary VL, which could indicate evolution of drug resistance during repeated treatment (Fig. 5). This comparison was, however, only significant at α=0.05 and could not be observed in longitudinal comparisons within a patient (Fig. S7). A larger sample size would be needed to support the role of this variant as an important factor in driving worse treatment outcomes in VL/HIV patients.

**Figure 5.**
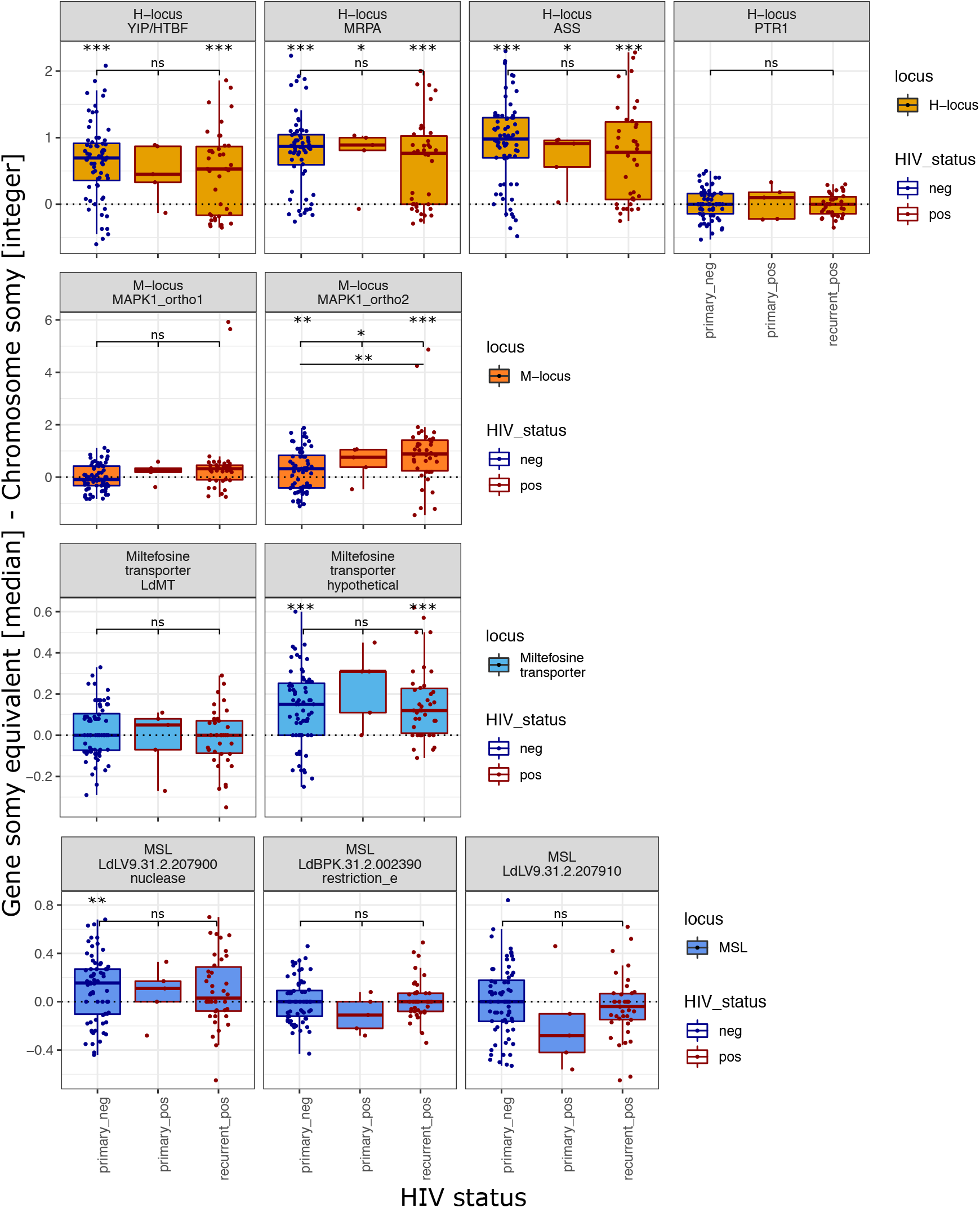
Gene copy numbers at known drug resistance loci across disease cohorts. Gene copy numbers at four drug resistance loci are shown for the three disease cohorts “primary_neg”, “primary_pos” and “recurrent_pos”. Gene copy numbers are estimated as differences in somy- equivalent to the respective chromosome somy. Upper row significance levels in each plot indicate FDR values of three One-sample T-Tests, in the second row ANOVA and in the third row TukeyHSD (if applicable) results. Significance levels are ***<0.001,*<0.01, *<0.05 and ns. not significant.

It has previously been suggested that different aneuploidy patterns themselves can be adaptive in new environments (Dumetz et al. 2017; Prieto Barja et al. 2017). We noticed that the dominant aneuploidy profile described above (all chromosomes are diploid except for a tetraploid chromosome 31) was more frequent in primary VL isolates before treatment (88.55%; 62/70) than in isolates from recurrent VL (44.7%; 17/38) (Fig. S8A). This poses the question of whether these different aneuploidy patterns might be adaptive in HIV patients, allowing parasites with these patterns to become more frequent in these patients during repeated VL episodes in HIV patients. We therefore tested if divergence from the dominant aneuploidy profile increased with the number episodes of VL relapse (Fig. S8C,D), but there is only support at α=0.1 for this trend (linear model: sum_diff_profile ∽ num_relapses, num_relapses p-value=0.0561). In total, 13 of 25 isolates from different patients with recurrent VL showed six different divergent aneuploidy profiles (Fig. S8B). In those profiles, a total of six chromosomes showed changes in somy in a few combinations. Four of the deviations were present in isolates from different patients with the most frequent changes occuring in 28% and 20% of all isolates from recurrent VL showing a reduction from a somy of 4 to 3 for chromosome 31 and a somy increase from 2 to 3 for chromosome 7 (Fig. S8B). While within patient-environments are expected to be heterogeneous due to both patient-specific differences and different drug treatment histories, this convergence might suggest an adaptive benefit of reduced chromosome 31 dosage or other convergent somy increases during prolonged parasite evolution in HIV patients with recurrent VL.

## Materials and Methods

This study was approved by the Institutional Review Board of the University of Gondar (IRB, reference O/V/P/RCS/05/1572/2017), the National Research Ethics Review Committee (NRERC, reference 310/130/2018) and Imperial College Research Ethics Committee (ICREC 17SM480). Informed written consent was obtained from each patient. The patients were all recruited at the time of VL diagnosis (ToD) at the Leishmaniasis Research and Treatment Center in Gondar, Amhara National Regional State, Ethiopia between December 2017 and May 2019 and were each followed up for three years (Takele et al. 2021).

### Parasite isolation and culturing

Isolates were obtained from splenic or bone marrow aspirates from VL and VL/HIV patients. Splenic aspiration can only be performed if the spleen is palpable at least 3 cm below the costal margin and there are no signs of bleeding, jaundice, severe anemia and platelet count < 40,000/ml. Otherwise, bone marrow aspiration was performed. Immediately after collection, tissue aspirations were cultured as previously described (Pescher et al. 2011) and kept in primary medium until DNA was extracted using DNeasy Blood & Tissue extraction Kits (QIAGEN, Hilden, Germany) according to the manufacturer’s instructions. Extracted DNA samples were stored at - 20°C before library preparation and sequencing.

### Sequencing

Genomic DNA was sheared into 400 to 600 base pair fragments by focused ultrasonication (Covaris Inc.), and Illumina libraries were prepared using NEB Ultra II custom kit (Kozarewa et al. 2009) then cleaned up using Agencourt AMPure XP SPRI beads (Beckman Coulter). The resulting libraries were sequenced as multiplexed pools of 96 samples and 17 samples (combined with samples from unrelated studies) as 151 bp paired-end reads on the Illumina Hiseq X10 platform. Sequencing reads from this project are all available from the European Nucleotide Archive as project PRJEB30077.

### Genomic analysis pipeline

Sequencing data for all samples were trimmed with Trimmomatic, version 0.39 (Bolger, Lohse, and Usadel 2014) to remove putative remains of adapter sequences and trim low quality 3’ end using parameters: “ILLUMINACLIP:PE_adaptors.fa:2:30:10 TRAILING:15 SLIDINGWINDOW:4:15 MINLEN:50” in paired-end mode. Resulting paired-end reads were mapped with BWA version 0.7.17 (Li and Durbin 2009) using the bwa mem -M option against the reference genome of the *L. donovani* isolate LV9 from Ethiopia, version 43 (https://tritrypdb.org/). SNPs were called on the resulting individual bam files using GATK version 4.1.2.0 (Poplin et al., n.d.; Auwera et al. 2013) with the Haplotype caller and parameters “-ERC GVCF --annotate-with-num-discovered-alleles --sample-ploidy 2” to generate gvcf files for each sample. “GenomicsDBImport” and “GenotypeGVCFs” were used to combine individual gvcf files and call SNPs for the sample cohort. SNPs were then hard filtered with “VariantFiltration” using the filters: QD < 2.0, MQ < 50.0, FS > 20.0, SOR > 2.5, BaseQRankSum <-3.1, ClippingRankSum <-3.1, MQRankSum <-3.1, ReadPosRankSum <-3.1and DP < 6”. For subsequent phylogenetic and population genetic analysis, SNPs with more than 20% missing calls across samples were removed, reducing the total number of SNPs to 370,714.

### Phylogenetic reconstruction and phenotype association

Phylogenetic trees were reconstructed using a distance based method: pairwise distances between samples were calculated as Nei’s genetic distances using the R package StAMPP version 1.6.1 (Pembleton, Cogan, and Forster 2013) and trees were reconstructed with neighbour joining using the R package ape version 5.4 (Paradis and Schliep 2019). To obtain bootstrap values, the total number of SNPs was drawn from all SNPs with replacement for each of 100 bootstrap replicates. For each, Nei’s distance matrices were calculated and individual replicate trees reconstructed with neighbour joining. Reported values at the nodes of the original tree are percentages of the respective node across the replicate trees. Associations of HIV and VL recurrence status with the phylogeny were tested based on the pairwise genetic distance matrix (Nei’s D) using ANOSIM analysis (R package vegan, version 2.5-6) and Blomberg’s *K* (R package phytools, version 0.7-47; Blomberg, Garland, and Ives 2003).

### Population genomics analysis

Population genomic analysis was based on genotype calls for each sample for the previously described 370714 SNP sites. All analysis and plotting was performed in R (R Core Team 2017). Alleles at all SNP sites were polarized based on their frequencies across all primary isolates and encoded as either H and L, standing for “high” and “low” frequency across all samples isolated from patients with primary VL. During time series analysis it was noticed that the two samples isolated from patient 1040 had unusually high fractions of LL genotypes during recurrent VL that was much higher than in isolated from other patients with recurrent disease (Fig. S9A). We therefore checked if this could be a biological signal or rather a technical artifact: Generally, high fractions of LL genotypes in a sample were associated with higher missingness of genotype calls (Fig. S9B) suggesting that extreme fraction of LL genotypes might rather be a genotype calling artifact in samples with less power of accurate SNP calling. We therefore excluded samples with a fraction of >= 0.002 unknown genotype calls from this analysis (Fig. S9C).

### Evaluation of complexity of infection

In a strictly clonal population allele frequency distributions have a clear expectation given the ploidy of the respective chromosome, e.g. for a diploid chromosome the allele frequency distribution should peak at 0.5 and for a triploid chromosome at ⅓ and/or ⅔. We used this expectation to evaluate whether each isolate was most likely to represent A) a single clonal infection where the allele frequencies were as expected, B) noisy profiles with very few variants or without very clear peaks or C) profiles with peaks at unexpected allele frequencies, likely to represent complex infections with a mixture of clones (Fig. S4A-C). For each isolate the allele frequency distribution for all variants called in that isolate on each diploid chromosome and the absolute difference between each allele frequency and the expected frequency of 0.5 was calculated. The distribution of these differences were summarised by its mean and standard deviation (Fig. S4D,E), confirming that complex infections were distinct from either single infection profiles or those with little, or noisy signal.

### Aneuploidy and copy number variant analysis

For copy number estimation previously described bam files were further processed to estimate somies of individual samples and chromosomes as well as the relative copy number of genes previously suggested to be involved in drug resistance. GATK, version 3.6, was used to perform indel realignment using the “RealignerTargetCreator” to identify and the “IndelRealigner” for realignment in the identified regions (Auwera et al. 2013). Then bam files were filtered for proper pair mapping and duplicates were removed using samtools, version 1.9 with the parameters “-F 1024 -f 0×0002 -F 0×0004 -F 0×0008”. Genome coverages were estimated with bedtools genomecov, version 2.29, and parameters “-d -split”. Data was subsequently processed in R. Aneuploidy profiles for each sample were estimated using median chromosome coverage. The median chromosome coverage for each sample was used to estimate the chromosome-specific coverage and across all chromosomes of a sample the median value was assumed to present the diploid stage. So for each sample: chromosome-specific somy = chromosome-specific coverage / median chromosome-specific coverage across all chromosomes * 2. Visualisations of aneuploidy profiles were done in R with the packages heatmap.2 and minimum spanning networks with the mst function of the pegas package (version 0.13).

To assess the coverage at genes of putative interest for drug resistance, coverage was estimated through the median coverage across bases in the respective gene. This coverage was translated into a somy-equivalent by dividing the base coverage at each genomic position by the median chromosome coverage and multiplying with the chromosome specific rounded somy. The somy equivalent change in coverage was estimated for genes from four loci previously associated with drug resistance including the H-locus, the M-locus (MAPK1) associated with resistance to antimonial drugs, the Miltefosine transporter and associated genes (Pérez-Victoria et al. 2006; Shaw et al. 2016) and the Miltefosine sensitivity locus. Genes in the LV9 reference genome (version 43) were identified via ortholog annotation in TriTryp (https://tritrypdb.org/) of the respective gene names of *L. infantum* JPCM5 (version 41) previously summarised in (Franssen et al. 2020); locus IDs for the LV9 genome annotation are included in figure 5.

## Discussion

We have shown that in Northern Ethiopia the same diverse population of parasites are responsible for VL and VL/HIV cases. Infection of HIV positive patients and the associated clinical manifestations (Takele 2021) is thus not due to phylogenetically distinct strains. However, we observe a small degree of genetic structure between isolates from VL and VL/HIV patients with borderline statistical significance. While we cannot exclude the possibility that specific groups of strains may have a higher propensity to infect HIV positive patients, we propose that this is likely to be due to geographic structure in the parasite population that is correlated with geographic differences in HIV prevalence. Many of the VL patients in this region of Ethiopia (Gadisa et al. 2015) – including 84.8% of the patients in this cohort (Takele et al. 2021) – are migrant workers who move from non-endemic regions for seasonal agricultural work. Possibly due to their non-endemic origin and the working and living conditions on the farms they are frequently infected with *Leishmania* and develop clinical VL. HIV infection is also more frequent in the migrant worker population (Tiruneh, Wasie, and Gonzalez 2015), introducing an association between the location of infection and HIV status. Identifying the actual location these patients acquired VL infections would require a detailed study based on these farms. These are extremely challenging environments to work in, both because of the lack of basic infrastructure and currently a poor security situation in the farming area close to the border with Sudan.

Our within-patient longitudinal data suggests that recurrent VL in HIV coinfected patients is predominantly caused by the re-expansion of persistent parasites. In six out of seven patients for which we could obtain longitudinal samples from subsequent relapses of VL, parasites from the same patient were clearly most closely related to each other. For one patient (1023), parasite isolates could be extracted at four different time points and show a gradual divergence from the initial isolate, as expected if parasites from the originally infecting population are responsible for relapses. In only a single case (patient 1045), isolates taken at the third and fourth episodes of VL for this patient (the time of initial diagnosis in this study and 7 months later) were as divergent as isolates from different patients. We interpret this as likely due to re-infection of this patient following cure. While we cannot rule out the possibility that this patient was initially infected with multiple parasite strains and by chance different strains from an initial infection would have been observed in subsequent isolates. We think this scenario is unlikely as we cannot see any strong indication that any isolates from this patient are complex infections (Fig. S5). We conclude that in this single case re-infection was contributing to the relapse of VL. The observation in the other six cases that parasite isolates from a single patient gradually diverge with the number of VL relapses, provides the first genetic evidence that successful treatment of VL – at least in HIV co-infected patients – is not sterile and is responsible for relapse.

It is clear from animal models of cutaneous leishmaniasis, that *Leishmania* can enter a distinct quiescent, non-replicating state (Mandell and Beverley 2017) that has been associated with persistence in tissues. Parasite persistence certainly also occurs clinically. Parasite DNA and live parasites have been isolated from healed CL scars (Mendonça et al. 2004) and from the skin of cured PKDL patients (Hossain et al. 2017). Re-appearance of active CL has been recorded up to 50 years after patients leave an endemic area (Czechowicz et al. 1999). This phenomenon also occurs with VL (Simon et al. 2011), sometimes with fatal consequences (Broeckaert-van Orshoven, Michielsen, and Vandepitte 1979). The presence of persistent parasites may be vital in establishing effective, long-term immunity to leishmania re-infection (Sacks 2014). The role of persistent parasites is also supported by our finding of much lower heterozygosity in parasite isolates from relapse episodes of VL, with a gradual reduction in heterozygosity with multiple relapses. Together, it suggests that heterozygosity in the infrapopulation of parasites within a host is lost due to strong population size reduction during initial VL treatment. In many HIV positive patients the population re-expands, causing relapse of disease symptoms: treatment of recurrent episodes of VL then leads to repeated population bottlenecks and continuing small reductions in heterozygosity. While this scenario is consistent with our data, a number of questions remain. We find a number of isolates that show unusual allele frequency distributions suggestive of complex infections caused by multiple clones, these were no more frequent in primary isolates than those taken during relapse episodes of VL, suggesting that loss of clonal complexity in the infections is not responsible for the observed loss of heterozygosity. We note that this is based on only a single isolate from spleen or bone marrow - it is possible that parasite populations are present in other host tissues such as the skin (Kirstein et al. 2017).

We find complex infections present as late as the tenth episode of VL relapse, which is very puzzling, although for these isolates the total heterozygosity is low that it might have been generated during within-host evolution. The origins of the heterozygosity present in the initial infection is not completely clear. One scenario could be the frequent presence of multiple infecting clones in the initial sandfly bite, most of which are lost on initial treatment. This would imply either that flies frequently pick-up multiclonal infections from single mammal hosts, or that they frequently bite multiple infected hosts before transmitting an infection. A few studies have either investigated multiple isolates collected from individual hosts (Cupolillo et al. 2020; Zackay et al. 2018), or directly investigated the diversity of parasites in tissues (Domagalska et al. 2019) or vectors (Parvizi et al. 2010; Darvishi et al. 2015) and find some evidence for polyclonal infections, but such reports seem to be rare. Similarly, individual sand flies biting more than two infected hosts seems unlikely to be very frequent when prevalence of *Leishmania* infections is not very high. However, it might be promoted if prevalence is much higher in very focal areas and because multiple blood meals appear to greatly promote *Leishmania* infections in the sandfly gut (Serafim et al. 2018). This could tend to increase the number of flies transmitting infections picked up from multiple hosts. In general, we know too little about the lifespan or biting rates of sandflies to evaluate this possibility, even in the best studied foci of VL (Cameron et al. 2016). Prevalence in this region is not well understood as quantitative data is either old (e.g. Fuller et al. 1976) or restricted to sub-populations such as HIV patients (e.g. van Griensven et al. 2019) or patients reporting with suspected VL (e.g. Ferede et al. 2017), although detailed information about one small neighbouring focus is available (Kirstein et al. 2018).

While to our knowledge we present the first longitudinal data from cases of *Leishmania* relapse, a number of studies have used genomic approaches to investigate recurrent malaria infections, which have demonstrated that relapse is due to parasite recrudescence (Rutledge et al. 2017; Guggisberg et al. 2016; Popovici et al. 2018). While these studies do not identify any genetic variation associated with relapse, they confirm that this is largely due to regrowth of one, or few, clonal lineages from initially complex infections (Rutledge et al. 2017; Popovici et al. 2018). One key limitation of our analysis of longitudinal changes in parasite populations is the ethical constraint on the availability of parasite isolates, which we could only obtain when invasive spleen or bone marrow aspirates were taken by clinicians for diagnostic purposes. Isolating parasites from blood samples would be less invasive (Dereure et al. 1998; Hide et al. 2007), but these approaches are little used and require conditions not generally available in resource-limited clinical settings. Culture-free approaches could be feasible (Domagalska et al. 2019), although parasitaemia in many VL patients is likely to be too low for current approaches (e.g. Sudarshan et al. 2015).

The heterozygosity we report in primary VL isolates appears to be due to both complex infections and genuine heterozygous variants within a parasite clone. These multiple causes of heterozygosity is one reason we have not attempted to use our diversity estimates to directly quantify the size of any population bottleneck in these patients. Another barrier to quantification is the rather complex genetics of *Leishmania. Leishmania* can reproduce clonally but also have normal, meiotic sex (Akopyants et al. 2009), which is probably rare (Rogers et al. 2014), and might also be able to reproduce parasexually (Sterkers et al. 2014). *Leishmania* populations also show extensive variation in aneuploidy (Downing et al. 2011; Mannaert et al. 2012; Sterkers et al. 2014; Imamura et al. 2016; Zackay et al. 2018; Patino et al. 2020; Franssen et al. 2020), but it is unclear to what extent this occurs in the field (Dumetz et al. 2017; Domagalska et al. 2019). Indeed, this aneuploidy variation seems to occur particularly rapidly in culture (Sterkers et al. 2012, 2014). Very rapid turnover of chromosome copies should decrease heterozygosity within individual cells, but it is not clear that it should impact heterozygosity measured at the level of an entire population of cells (Sterkers et al. 2014). Robust approaches to estimate parameters such as the change in population size upon drug treatment from patterns of genetic variation between isolates or through time in a longitudinal sample would need to take these unusual features of *Leishmania* genetics into account.

We report a relatively small amount of aneuploidy variation between isolates, which could contribute to the loss of heterozygosity we observe with treatment of VL relapse.. An important caveat to these results is that we estimate ploidy from DNA extracted from the entire population of cells present in the primary taken to isolate parasites. Using these initial cultures minimises the possibility that aneuploidy changes occur as the parasites adapt to *in vitro* growth, but there could still be some difference to the aneuploidy profile of parasites in the patients themselves (Domagalska et al. 2019; Dumetz et al. 2017). In addition, the measure will be conservative in identifying variation in somy, as it ignores variation between parasite cells within a single isolate. Directly demonstrating that aneuploidy turnover occurs and quantifying its impact on heterozygosity differences between isolates would require either multiple samples from a much larger set of patients, or investigating the genomes of individual cells directly, which is now possible but remains costly, technically challenging and gives much less clear results than sequencing larger amounts of material (Imamura et al. 2020)

The parasites we investigate from recurrent VL cases have not been phenotypically investigated for drug sensitivity or resistance, and we do not generally identify drug resistance in the genomic data from these parasites. We thus presume that drug resistance is not responsible for the increased difficulty in treating patients with repeated relapses of VL (Mohammed et al. 2020; Takele et al. 2021) and the failure to successfully treat these patients long term. Indeed, it is unclear in general to what extent treatment failure in *Leishmania* is due to drug resistance (Yardley et al. 2006; Rijal et al. 2007; Ponte-Sucre et al. 2017). However, we can only screen for known genetic variants associated with resistance, and our knowledge of the genetic basis of drug resistance in *Leishmania* is very incomplete (Ponte-Sucre et al. 2017) and largely based on studying parasite clones selected for resistance in the laboratory (Croft, Sundar, and Fairlamb 2006). In particular, the markers that are available for VL have largely been identified in Indian *L. donovani* or in *L. infantum*. African *L. donovani* populations are genetically highly differentiated from these populations elsewhere in the world (Franssen et al. 2020) and it is possible that the genetic basis of drug resistance in Africa, if any, could be very different. In particular, many known markers are for resistance to miltefosine, which is not widely available in Africa (Sunyoto, Potet, and Boelaert 2018). Little work has been done to investigate drug resistance markers in African visceral leishmaniasis, where drug resistance is not thought to be a significant clinical problem (Gidey et al. 2019), and no genetic markers have – to our knowledge – been identified in the region. Identifying such resistance in recurrent VL/HIV would be complicated by the complex history of drug exposure for many of these patients, as clinicians frequently resort to second-line drugs or combinations in an attempt to improve treatment outcomes (Diro et al. 2019; Monge-Maillo and López-Vélez 2016; Mohammed et al. 2020).

## Conclusion

We show that a diverse population of *L. donovani* present in Northern Ethiopia is responsible for visceral leishmaniasis in both single VL infections and in VL/HIV co-infections, with no clear genome-wide genetic differentiation between parasites isolated from the two patient groups. Our data suggest that most relapses are caused by recrudescence (regrowth) of the initial infecting parasite population, rather than re-infection from a subsequent infected sandfly bite. Finally, we show that the parasite populations present in primary VL episodes are more diverse than those isolated during VL relapses, and that the infrapopulation of parasites within a host patient loses genetic diversity with increasing numbers of subsequent relapses. This loss of diversity is presumably due to population bottlenecks induced by treatment of each episode of disease. We observe complex multi-clonal infections but these are not more common in primary infections. We propose that infecting parasites are able to re-establish despite clinical cure after anti-leishmanial treatment; due to the dysfunctioning immune system of patients coinfected with VL and HIV (Takele et al. 2021) that is unable to prevent re-expansion of the parasite infrapopulation leading to VL relapse. This implies that better approaches to identifying patients at highest risk of relapse (Takele et al. 2021), and alternative treatment that allows these patients to restore functioning immunity might help prevent subsequent relapses and so improve clinical outcomes for VL/HIV patients.

## Data Availability

All data is available at the ENA as project PRJEB 30077, as listed in table S1.

## Acknowledgements

This work was supported by Wellcome via a Training Fellowship in Public Health and Tropical Medicine to YT (grant 204797/Z/16/Z) and core support from the Wellcome Sanger Institute (WT206194) and jointly by the UK Medical Research Council (MRC) and the UK Department for International Development (DFID) under the MRC/DFID Concordat agreement via grants MR/R01020X/1 and MR/R021600/1. We acknowledge and thank all members of the study teams from Ethiopia, the Leishmaniasis Research and Treatment Centre of the University of Gondar.

## Notes

### Competing Interest Statement

The authors have declared no competing interest.

### Author Declarations

Institutional Review Board of the University of Gondar (IRB, reference O/V/P/RCS/05/1572/2017), the National Research Ethics Review Committee (NRERC, reference 310/130/2018) and Imperial College Research Ethics Committee (ICREC 17SM480).

