## Supplementary Information for "Diversity and within-host evolution of parasites from VL and VL/HIV patients in Northern Ethiopia"

### Table of contents

**Fig. S1 Genomic relatedness between first parasite isolates taken from each patient of primary VL only.** 2

**Fig. S2 Heatmap of Nei's distances for isolates of patients 1023 and 1045.** 3

**Fig. S3 Relationship of the heterozygous fraction with other disease phenotypes.** 4

**Fig. S4 Use of allele frequency estimates to evaluate clonal diversity of individual isolates.** 5

**Fig. S5 Allele frequency profiles for isolates from patient 1045.** 7

**Fig. S6 Aneuploidy profiles of all 113 parasite samples.** 8

**Fig. S7 Gene copy numbers at known drug resistance loci for time series isolates.** 10

**Fig. S8 Aneuploidy changes of parasites from patients with recurrent VL.** 11

**Fig. S9 Samples excluded from heterozygosity analysis due to potential quality issues.** 13

**Table S1: Summary of all isolates in this study.** 14

**Table S2: Aneuploidy profiles and metadata summary of all isolates from patients with time series data and /or replicates of primary isolates.** 21

Fig. S1 Genomic relatedness between first parasite isolates taken from each patient of primary VL only.

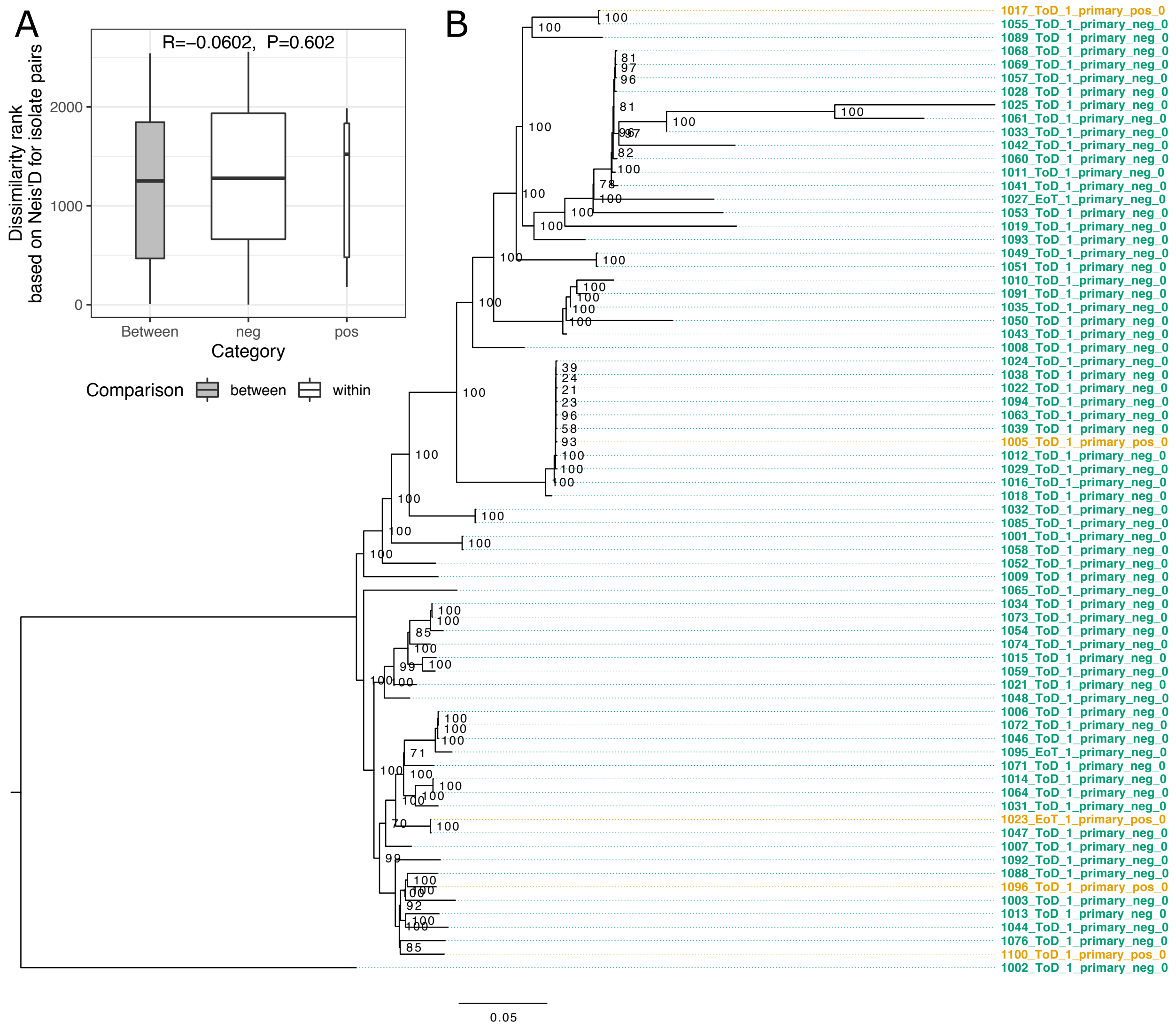


**Figure S1**. Genomic relatedness between first parasite isolates taken from each patient of primary VL only. A) ANOSIM results comparing sum of ranked pairwise genetic distances (Nei's D) within and between HIV positive and negative samples from primary VL only. B) Phylogeny of first isolate of primary infection taken from each patient. Sample colour indicates HIV status: green for isolates from HIV negative patients, light orange for HIV positive. Sample names are composed as in figure 1. Bootstrap values are indicated at branch nodes. The phylogeny is rooted based on the inclusion of an *L. infantum* outgroup (data not shown).

Fig. S2 Heatmap of Nei's distances for isolates of patients 1023 and 1045.

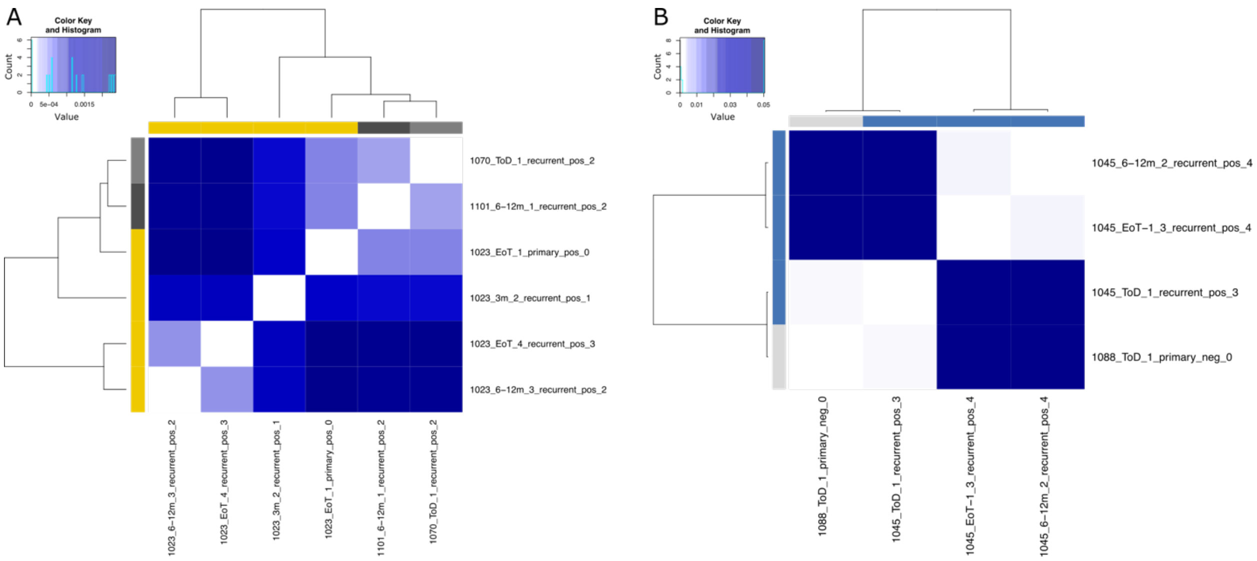


**Figure S2.** Heatmap of Nei's distances for isolates of patients 1023 and 1045. Heatmaps display pairwise distances between all samples of the respective patient from different time points of isolation and the closest sample from our entire sample collection if closer than the remaining isolates from the same patient. A) Four samples from patient 1023 isolated from primary VL and recurrent VL relapses 1, 2 and 3 are shown along with two isolates from different patients closest to the isolate from primary VL. Time series isolates diverge gradually with time. B) Three samples from patient 1045 were isolated at recurrent VL "relapse 3" and "relapse 4" before and after treatment. The isolate from recurrent VL "relapse 3" was closest to an isolate from another patient and is very different from the two isolates from the subsequent recurrent VL relapse.

Fig. S3 Relationship of the heterozygous fraction with other disease phenotypes.

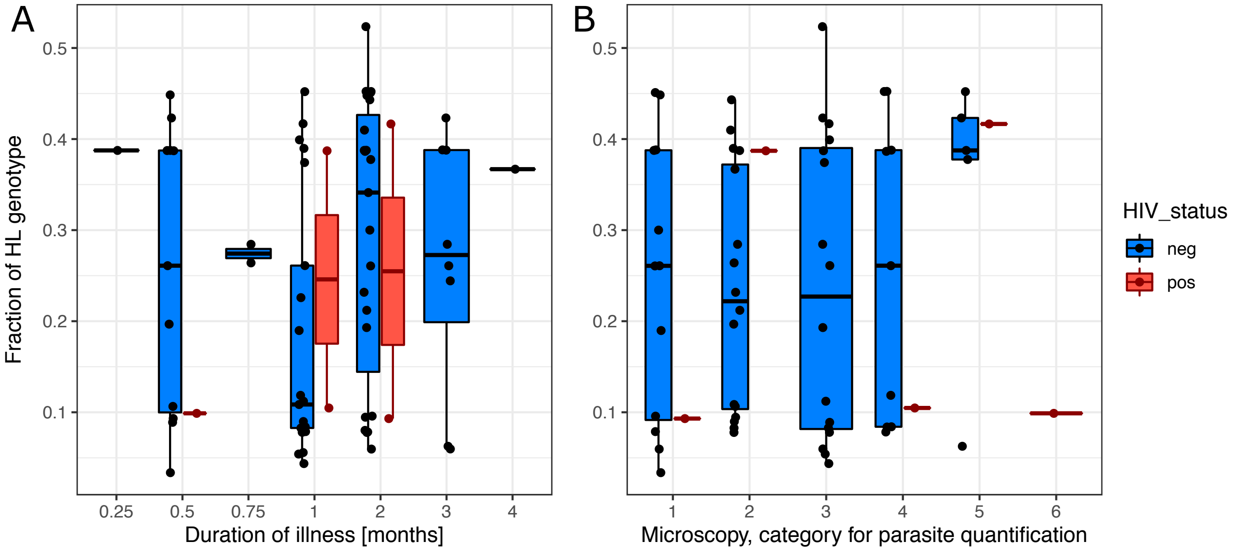


**Figure S3.** Relationship of the heterozygous fraction with other disease phenotypes. The fraction of heterozygotes in an isolate with respect to the A) duration of symptomatic primary VL and B) parasite load at the time of hospital acception for symptomatic primary VL. Linear models for both phenotypes of interest did not show any linear relationship with duration of illness or parasite load, respectively. Neither was HIV status an explanatory factor in either case. A) lm(fHL ~ duration_illness_months + HIV_status, duration_illness_months: estimate=0.0305, p-value=0.151, HIV_status: estimate=-0.0231, p-value=0.741) B) lm(fHL ~ microscopy + HIV_status, microscopy: estimate=0.0127, p-value=0.403, HIV_status: estimate=-0.0420, p-value=0.565).

Fig. S4 Use of allele frequency estimates to evaluate clonal diversity of individual isolates.

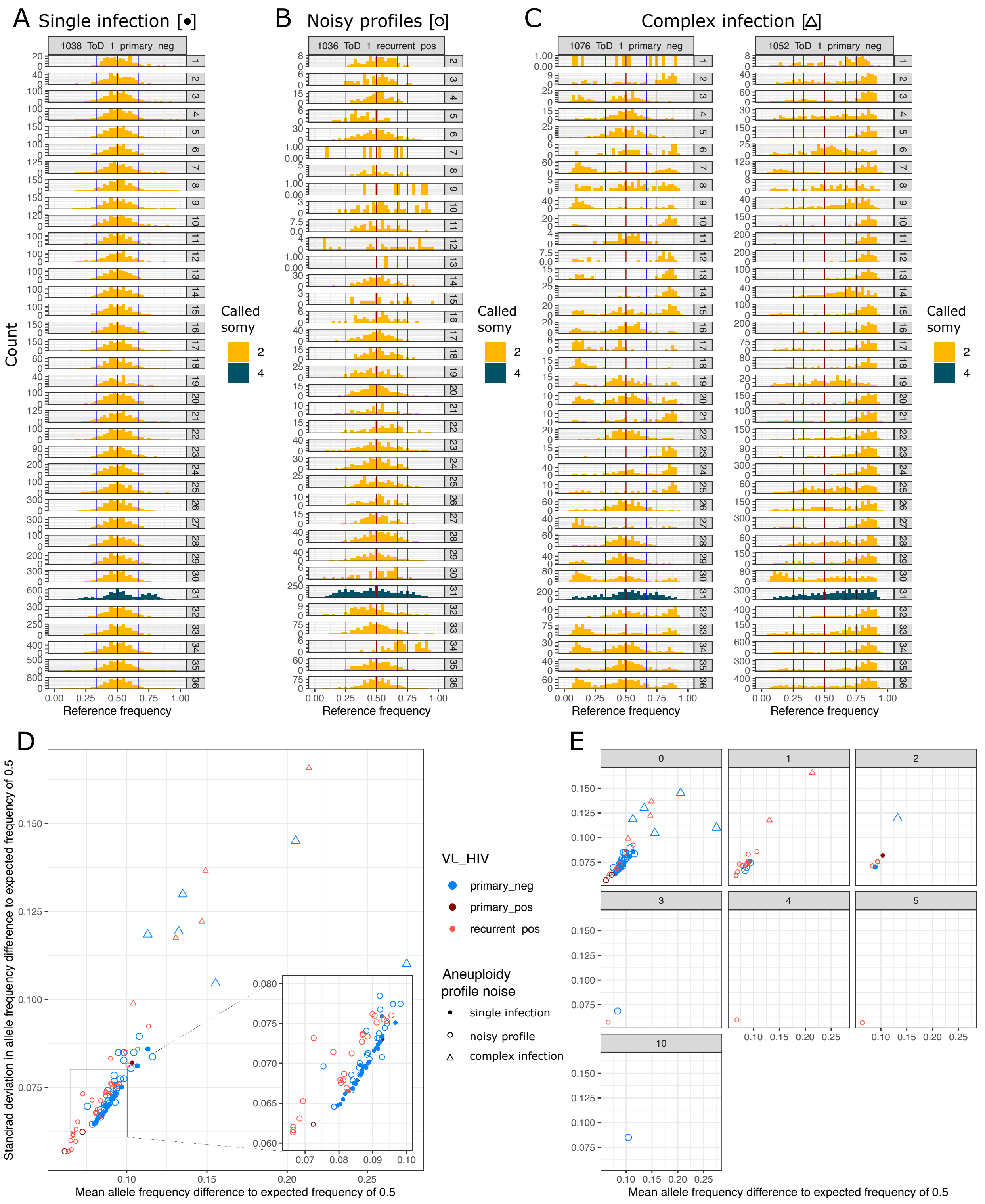


**Figure S4.** Use of allele frequency estimates to evaluate clonal diversity of individual isolates. Plots A-C display one or two examples of categories for the noisiness of aneuploidy profiles based on allele frequency estimates for each isolate. Categories were determined by visual inspection of the allele frequency plots by chromosome for each isolate. A) Single infection: allele frequency distributions across chromosomes peak at the frequency expected based on the chromosome somy without outliers from the distribution around the peak. B) Noisy profiles: while allele frequency distributions mainly peak around the expected mean, some noise is visible. C) Complex infection: allele frequency distributions have multiple peaks, also at high and low frequency. D) Mean and standard deviation (sd) across isolates from expected allele frequency of 0.5 for diploid chromosomes. Symbols are coloured by VL/HIV status. Symbol types display the category of the noise of the aneuploidy profile as shown in A-C. The visual categorisation is partly mirrored by the mean and sd of the deviation from the allele frequency expectation. E) Data from A is shown in seven different subplots that indicate the summed difference of the isolate's aneuploidy profile from the common aneuploidy profile (diploid for all chromosomes except for a tetraploid chromosome 31). Summed up differences are indicated at the top of each subplot.

Fig. S5 Allele frequency profiles for isolates from patient 1045.

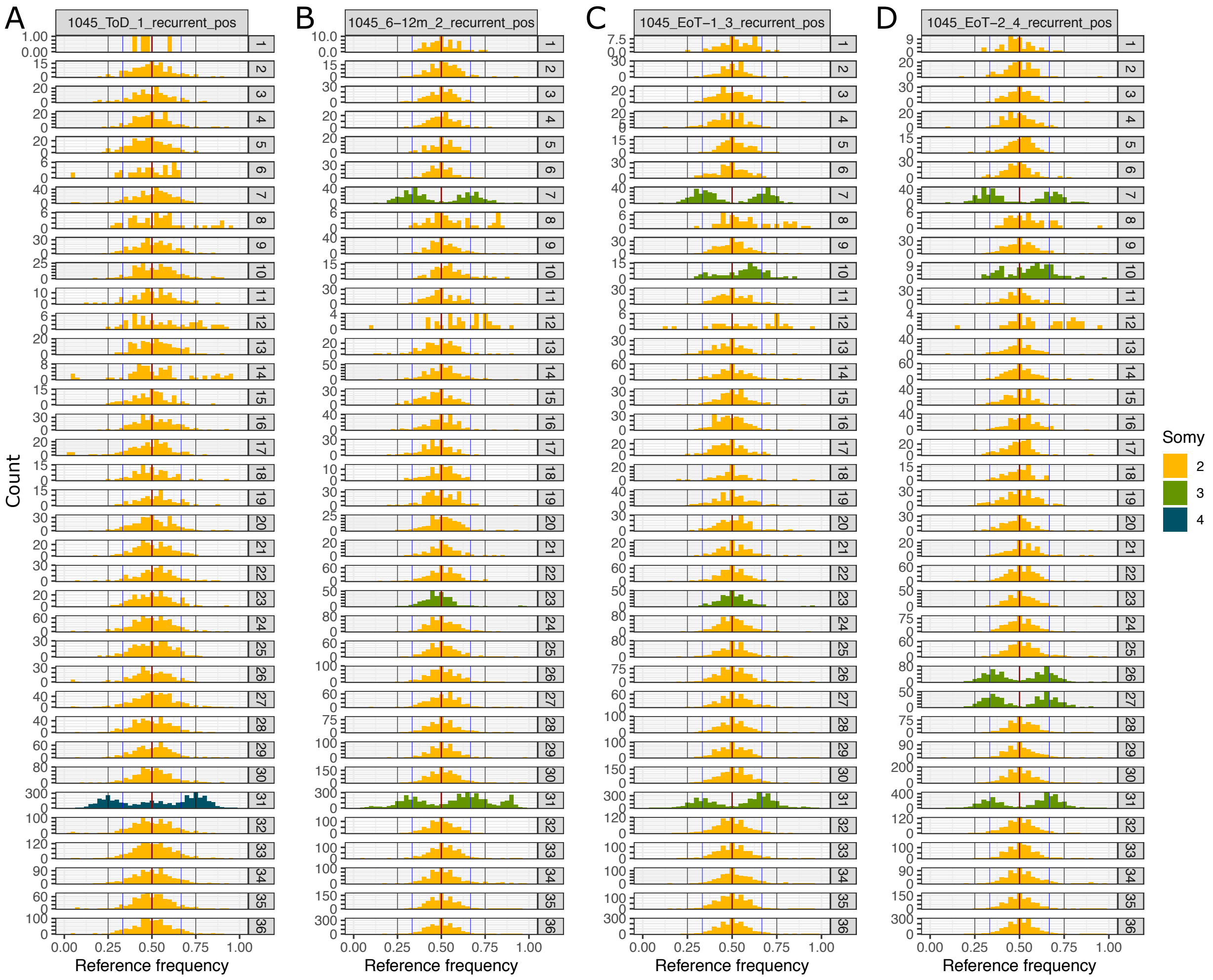


**Figure S5.** Allele frequency profiles for isolates from patient 1045. Allele frequency profiles are shown separately for each chromosome and isolate and color coded by the respective chromosome copy number. All profiles have been categorised as noisy though they dominantly follow an expected frequency distribution based on respective somies (Fig. S4).

Fig. S6 Aneuploidy profiles of all 113 parasite samples.

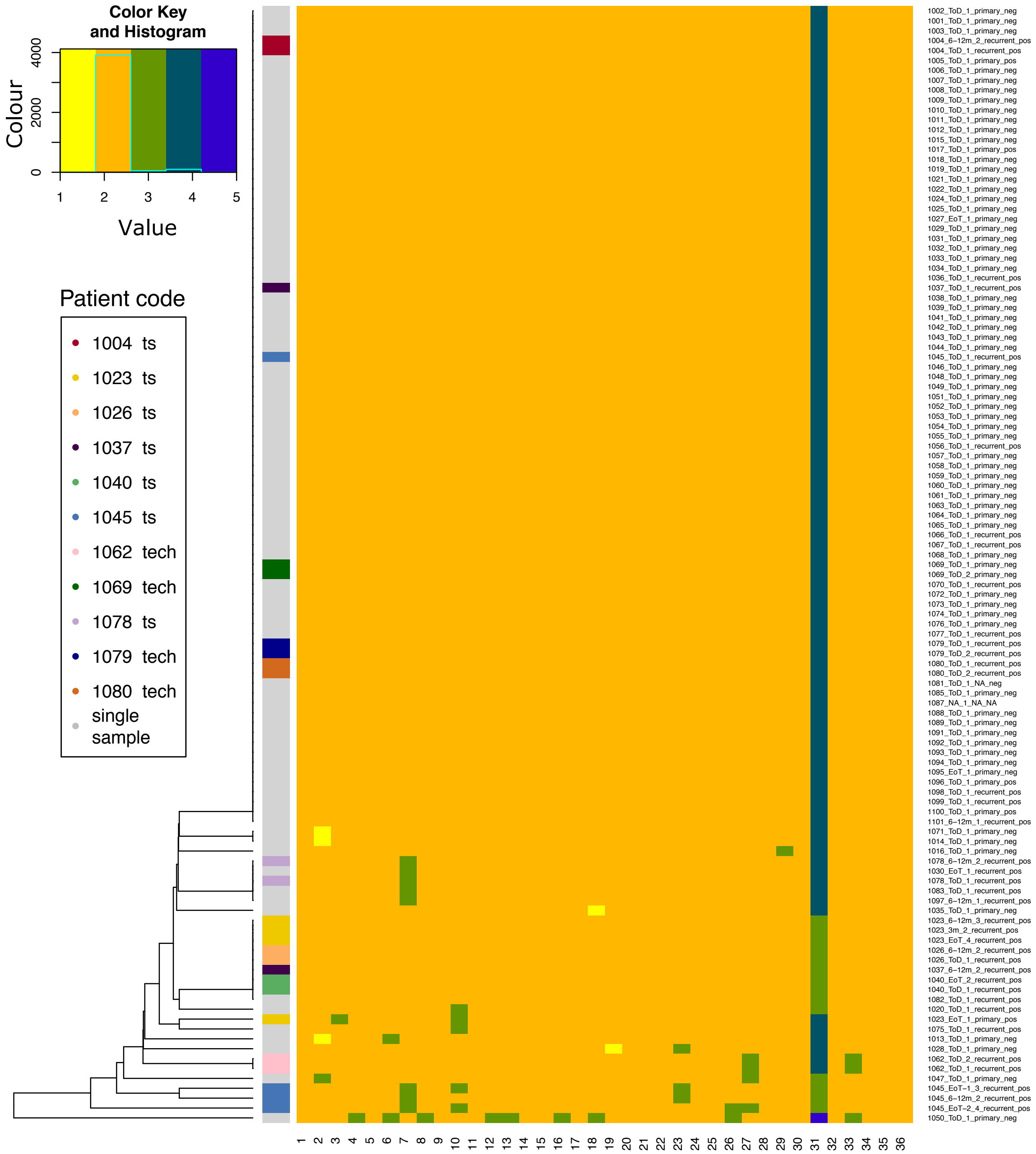


**Figure S6.** Aneuploidy profiles of all 113 parasite samples. The heatmap displays aneuploidy profiles with color coded somies for each patient (rows) and chromosome (columns). Sample identifiers are listed on the right-hand side of the respective row. The leftmost colour column indicates patient association of patients with time series data (ts) and /or isolate aliquots (tech) using different colors for each patient. Samples from all remaining patients with a single isolate only are coloured in gray. Rows with aneuploidy profiles are ordered with average linkage clustering with the cladogram shown on the left.

Fig. S7 Gene copy numbers at known drug resistance loci for time series isolates.

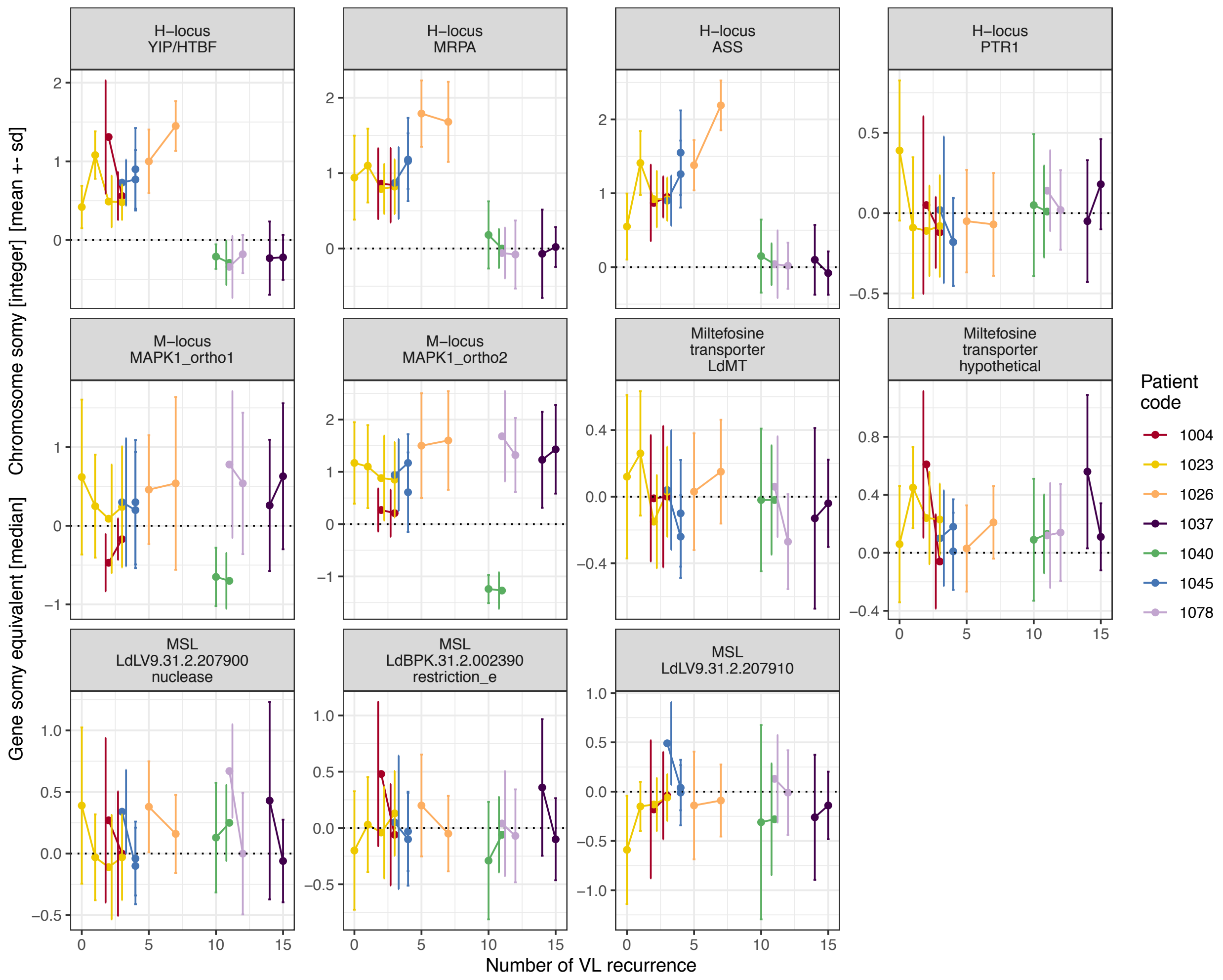


**Figure S7.** Gene copy numbers at known drug resistance loci for time series isolates. Gene copy numbers at drug resistance loci are shown for all the seven patients from whom we have multiple isolates sampled at different time points. Time is measured as the number of the recurrences VL episode, when the isolate was taken (0 is primary VL). Gene copy numbers are estimated as differences in somy-equivalent to the respective chromosome somy and shown are mean +/- standard deviation using individual base pair coverages within each gene.

Fig. S8 Aneuploidy changes of parasites from patients with recurrent VL.

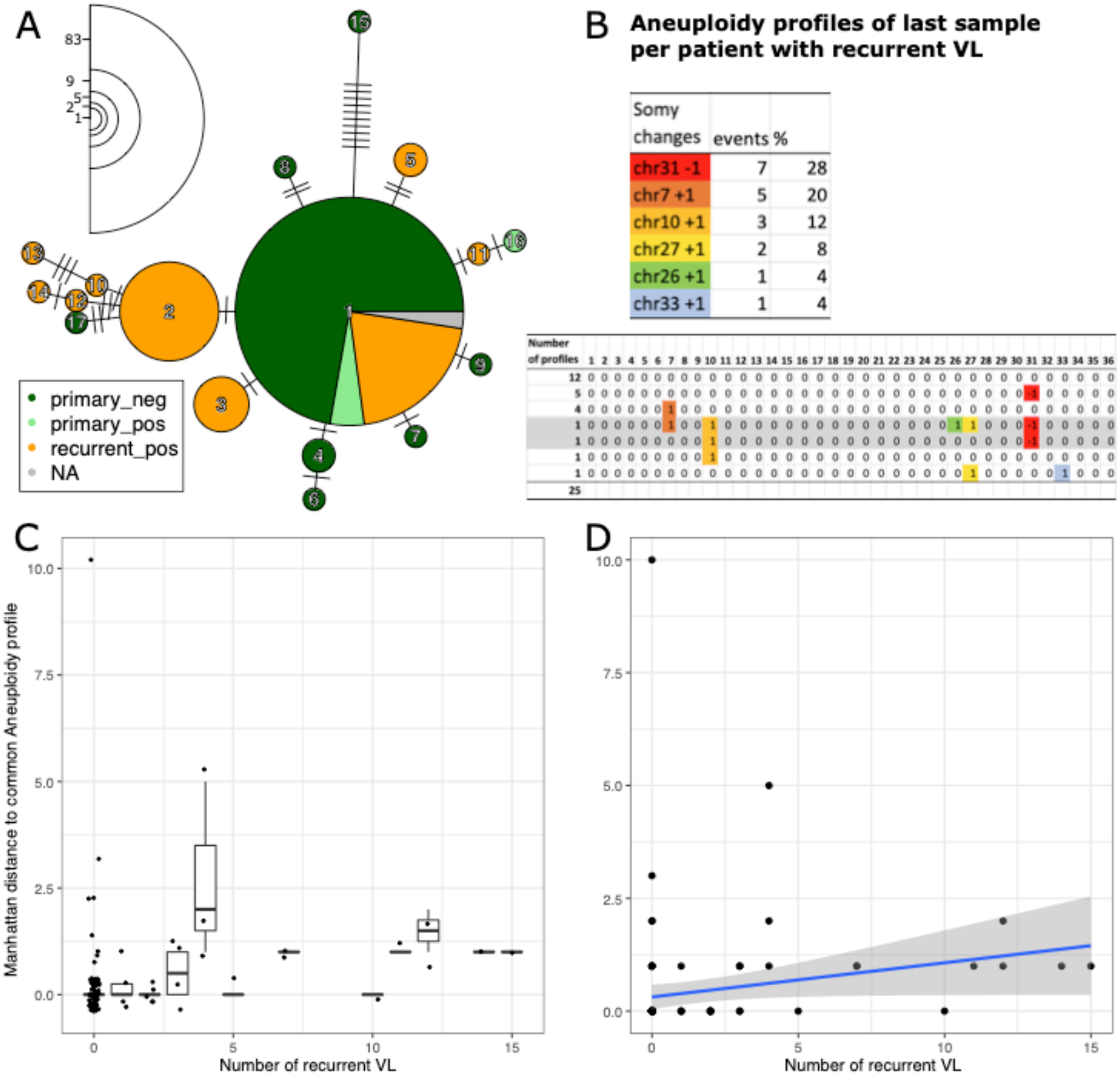


| Somy changes | events | % |
|---|---|---|
| chr31 -1 | 7 | 28 |
| chr7 +1 | 5 | 20 |
| chr10 +1 | 3 | 12 |
| chr27 +1 | 2 | 8 |
| chr26 +1 | 1 | 4 |
| chr33 +1 | 1 | 4 |

| Number of profiles | 1 | 2 | 3 | 4 | 5 | 6 | 7 | 8 | 9 | 10 | 11 | 12 | 13 | 14 | 15 | 16 | 17 | 18 | 19 | 20 | 21 | 22 | 23 | 24 | 25 | 26 | 27 | 28 | 29 | 30 | 31 | 32 | 33 | 34 | 35 | 36 |
|---|---|---|---|---|---|---|---|---|---|---|---|---|---|---|---|---|---|---|---|---|---|---|---|---|---|---|---|---|---|---|---|---|---|---|---|---|
| 12 | 0 | 0 | 0 | 0 | 0 | 0 | 0 | 0 | 0 | 0 | 0 | 0 | 0 | 0 | 0 | 0 | 0 | 0 | 0 | 0 | 0 | 0 | 0 | 0 | 0 | 0 | 0 | 0 | 0 | 0 | 0 | 0 | 0 | 0 | 0 | 0 |
| 5 | 0 | 0 | 0 | 0 | 0 | 0 | 0 | 0 | 0 | 0 | 0 | 0 | 0 | 0 | 0 | 0 | 0 | 0 | 0 | 0 | 0 | 0 | 0 | 0 | 0 | 0 | 0 | 0 | 0 | 0 | -1 | 0 | 0 | 0 | 0 | 0 |
| 4 | 0 | 0 | 0 | 0 | 0 | 0 | 1 | 0 | 0 | 0 | 0 | 0 | 0 | 0 | 0 | 0 | 0 | 0 | 0 | 0 | 0 | 0 | 0 | 0 | 0 | 0 | 0 | 0 | 0 | 0 | 0 | 0 | 0 | 0 | 0 | 0 |
| 1 | 0 | 0 | 0 | 0 | 0 | 0 | 1 | 0 | 0 | 1 | 0 | 0 | 0 | 0 | 0 | 0 | 0 | 0 | 0 | 0 | 0 | 0 | 0 | 0 | 0 | 1 | 1 | 0 | 0 | 0 | -1 | 0 | 0 | 0 | 0 | 0 |
| 1 | 0 | 0 | 0 | 0 | 0 | 0 | 0 | 0 | 0 | 1 | 0 | 0 | 0 | 0 | 0 | 0 | 0 | 0 | 0 | 0 | 0 | 0 | 0 | 0 | 0 | 0 | 0 | 0 | 0 | 0 | -1 | 0 | 0 | 0 | 0 | 0 |
| 1 | 0 | 0 | 0 | 0 | 0 | 0 | 0 | 0 | 0 | 1 | 0 | 0 | 0 | 0 | 0 | 0 | 0 | 0 | 0 | 0 | 0 | 0 | 0 | 0 | 0 | 0 | 0 | 0 | 0 | 0 | 0 | 0 | 0 | 0 | 0 | 0 |
| 1 | 0 | 0 | 0 | 0 | 0 | 0 | 0 | 0 | 0 | 0 | 0 | 0 | 0 | 0 | 0 | 0 | 0 | 0 | 0 | 0 | 0 | 0 | 0 | 0 | 0 | 0 | 1 | 0 | 0 | 0 | 0 | 0 | 1 | 0 | 0 | 0 |
| 25 | | | | | | | | | | | | | | | | | | | | | | | | | | | | | | | | | | | | |

**Figure S8.** Aneuploidy changes of parasites from patients with recurrent VL. A) Aneuploidy profiles of all 113 parasite isolates are shown in a minimum spanning network as in figure 4 but labeled by VL and HIV coinfection status. Connections indicate profile similarity with the number of ticks indicating the sum of somy differences between connected profiles. Circle sizes represent the numbers of isolates showing a particular profile and numbers for each circle size are indicated in the left upper corner. Circles are numbered to identify each aneuploidy profile, and details of the somy patterns and abundance of each profile are listed in table 2. The largest circle (profile 1) represents the diploid condition, but with a tetrasomic chromosome 31. B) Aneuploidy profiles of the last isolate taken from each patient with recurrent VL are summarised. The first table summarises the chromosomes changed with the type of change and the frequency of occurrence out of all 25 last samples per patient with recurrent VL. In the second table the respective profiles and number of occurrences are listed. C) & D) For aneuploidy profiles of all 96 last isolates from each patient with known relapse status Manhattan distances to the most common aneuploidy profile were calculated and shown with respect to the number of VL relapse isolated from. C) Boxplots with individual data points as jitter. D) Scatter plot with linear regression line and 95% confidence intervals.

Fig. S9 Samples excluded from heterozygosity analysis due to potential quality issues.

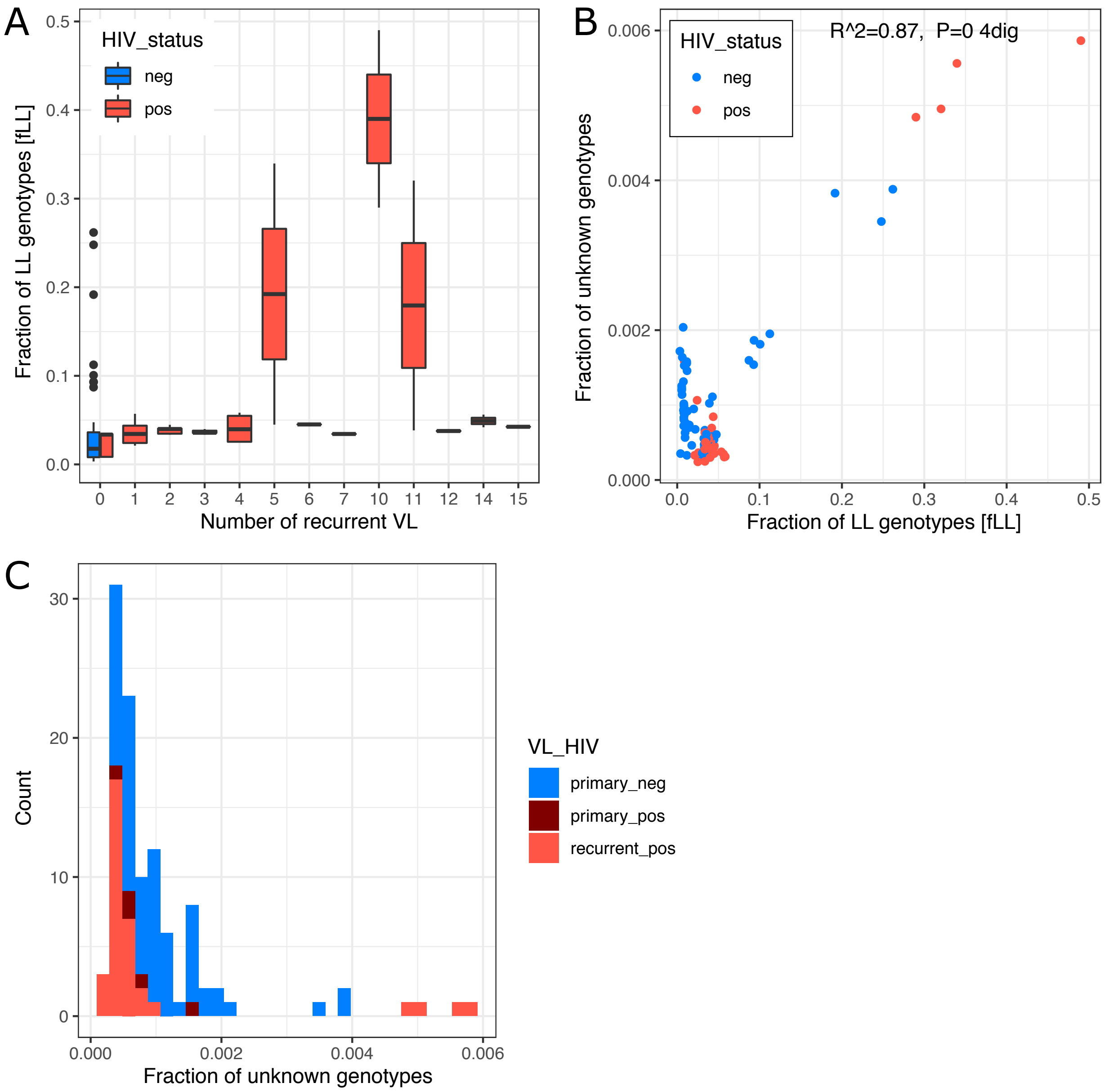


**Figure S9.** Samples excluded from heterozygosity analysis due to potential quality issues. A) Fraction of LL genotypes (L alleles are the low frequency allele across all isolates from primary VL) with respect to the VL episode of the patient isolated from. B) Correlation of the fraction of LL genotypes per isolate with fraction of unknown genotypes in that sample. C) Distribution of fraction of unknown genotypes across samples categorised by VL/HIV coinfection status. The sample names with fractions of unknown genotypes >= 0.002 are: 1002_ToD_1_primary_neg, 1025_ToD_1_primary_neg, 1040_EoT_2_recurrent_pos, 1040_ToD_1_recurrent_pos, 1041_ToD_1_primary_neg, 1061_ToD_1_primary_neg, 1077_ToD_1_recurrent_pos, 1079_ToD_1_recurrent_pos.

**Table S1.** Summary of all isolates in this study.

| Patient code | Sample ID | Date of disease within study | Site of parasite isolation | VL diagnosis | HIV status | Number of relapse | Microscopy category measuring parasite load | Duration of illness of current disease episode until hospitalisation [months] | Time series data available for patient | Only used as additional replicate for this sample | ENA study accession |
|---|---|---|---|---|---|---|---|---|---|---|---|
| 1001 | 1001_ToD_1_primary_neg | ToD | BM | primary | neg | 0 | 1+ | 2 | FALSE | FALSE | PRJEB30077 |
| 1002 | 1002_ToD_1_primary_neg | ToD | BM | primary | neg | 0 | 3+ | 0.5 | FALSE | FALSE | PRJEB30077 |
| 1003 | 1003_ToD_1_primary_neg | ToD | Spleen | primary | neg | 0 | 4+ | 1 | FALSE | FALSE | PRJEB30077 |
| 1004 | 1004_ToD_1_recurrent_pos | ToD | Spleen | recurrent | pos | 2 | 1+ | 2 | TRUE | FALSE | PRJEB30077 |
| 1004 | 1004_6-12m_2_recurrent_pos | 6-12m | Spleen | recurrent | pos | 3 | 1+ | 2 | TRUE | FALSE | PRJEB30077 |
| 1005 | 1005_ToD_1_primary_pos | ToD | BM | primary | pos | 0 | 2+ | 1 | FALSE | FALSE | PRJEB30077 |
| 1006 | 1006_ToD_1_primary_neg | ToD | Spleen | primary | neg | 0 | 2+ | 2 | FALSE | FALSE | PRJEB30077 |
| 1007 | 1007_ToD_1_primary_neg | ToD | Spleen | primary | neg | 0 | 3+ | 1 | FALSE | FALSE | PRJEB30077 |
| 1008 | 1008_ToD_1_primary_neg | ToD | Spleen | primary | neg | 0 | 1+ | 2 | FALSE | FALSE | PRJEB30077 |
| 1009 | 1009_ToD_1_primary_neg | ToD | Spleen | primary | neg | 0 | 2+ | 2 | FALSE | FALSE | PRJEB30077 |
| 1010 | 1010_ToD_1_primary_neg | ToD | Spleen | primary | neg | 0 | -ve | 2 | FALSE | FALSE | PRJEB30077 |
| 1011 | 1011_ToD_1_primary_neg | ToD | Spleen | primary | neg | 0 | 1+ | 0.5 | FALSE | FALSE | PRJEB30077 |
| 1012 | 1012_ToD_1_primary_neg | ToD | BM | primary | neg | 0 | 3+ | 0.5 | FALSE | FALSE | PRJEB30077 |

| | | | | | | | | | | | |
|---|---|---|---|---|---|---|---|---|---|---|---|
| 1013 | 1013_ToD_1_primary_neg | ToD | Spleen | primary | neg | 0 | 2+ | 0.5 | FALSE | FALSE | PRJEB30077 |
| 1014 | 1014_ToD_1_primary_neg | ToD | Spleen | primary | neg | 0 | 2+ | 1 | FALSE | FALSE | PRJEB30077 |
| 1015 | 1015_ToD_1_primary_neg | ToD | Spleen | primary | neg | 0 | 3+ | 1 | FALSE | FALSE | PRJEB30077 |
| 1016 | 1016_ToD_1_primary_neg | ToD | Spleen | primary | neg | 0 | 2+ | 1 | FALSE | FALSE | PRJEB30077 |
| 1017 | 1017_ToD_1_primary_pos | ToD | Spleen | primary | pos | 0 | 5+ | 2 | FALSE | FALSE | PRJEB30077 |
| 1018 | 1018_ToD_1_primary_neg | ToD | Spleen | primary | neg | 0 | 2+ | 4 | FALSE | FALSE | PRJEB30077 |
| 1019 | 1019_ToD_1_primary_neg | ToD | Spleen | primary | neg | 0 | 3+ | 2 | FALSE | FALSE | PRJEB30077 |
| 1020 | 1020_ToD_1_recurrent_pos | ToD | Spleen | recurrent | pos | 12 | 5+ | 1 | FALSE | FALSE | PRJEB30077 |
| 1021 | 1021_ToD_1_primary_neg | ToD | Spleen | primary | neg | 0 | 2+ | 1 | FALSE | FALSE | PRJEB30077 |
| 1022 | 1022_ToD_1_primary_neg | ToD | Spleen | primary | neg | 0 | -ve | 2 | FALSE | FALSE | PRJEB30077 |
| 1023 | 1023_EoT_1_primary_pos | EoT | BM | primary | pos | 0 | 1+ | 2 | TRUE | FALSE | PRJEB30077 |
| 1023 | 1023_3m_2_recurrent_pos | 3m | Spleen | recurrent | pos | 1 | 5+ | 2 | TRUE | FALSE | PRJEB30077 |
| 1023 | 1023_6-12m_3_recurrent_pos | 6-12m | Spleen | recurrent | pos | 2 | 6+ | 2 | TRUE | FALSE | PRJEB30077 |
| 1023 | 1023_EoT_4_recurrent_pos | EoT | Spleen | recurrent | pos | 3 | 2+ | 2 | TRUE | FALSE | PRJEB30077 |
| 1024 | 1024_ToD_1_primary_neg | ToD | Spleen | primary | neg | 0 | 1+ | 2 | FALSE | FALSE | PRJEB30077 |
| 1025 | 1025_ToD_1_primary_neg | ToD | Spleen | primary | neg | 0 | 1+ | 1 | FALSE | FALSE | PRJEB30077 |
| 1026 | 1026_ToD_1_recurrent_pos | ToD | Spleen | recurrent | pos | 5 | 5+ | 2 | TRUE | FALSE | PRJEB30077 |

| | | | | | | | | | | | |
|---|---|---|---|---|---|---|---|---|---|---|---|
| 1026 | 1026_6-12m_2_recurrent_pos | 6-12m | Spleen | recurrent | pos | 7 | 5+ | 2 | TRUE | FALSE | PRJEB30077 |
| 1027 | 1027_EoT_1_primary_neg | EoT | Spleen | primary | neg | 0 | -ve | 3 | FALSE | FALSE | PRJEB30077 |
| 1028 | 1028_ToD_1_primary_neg | ToD | BM | primary | neg | 0 | 1+ | 2 | FALSE | FALSE | PRJEB30077 |
| 1029 | 1029_ToD_1_primary_neg | ToD | Spleen | primary | neg | 0 | 4+ | 2 | FALSE | FALSE | PRJEB30077 |
| 1030 | 1030_EoT_1_recurrent_pos | EoT | Spleen | recurrent | pos | 7 | 5+ | 2 | FALSE | FALSE | PRJEB30077 |
| 1031 | 1031_ToD_1_primary_neg | ToD | BM | primary | neg | 0 | 4+ | 2 | FALSE | FALSE | PRJEB30077 |
| 1032 | 1032_ToD_1_primary_neg | ToD | Spleen | primary | neg | 0 | 1+ | 3 | FALSE | FALSE | PRJEB30077 |
| 1033 | 1033_ToD_1_primary_neg | ToD | BM | primary | neg | 0 | 3+ | 2 | FALSE | FALSE | PRJEB30077 |
| 1034 | 1034_ToD_1_primary_neg | ToD | BM | primary | neg | 0 | 3+ | 3 | FALSE | FALSE | PRJEB30077 |
| 1035 | 1035_ToD_1_primary_neg | ToD | Spleen | primary | neg | 0 | -ve | 0.5 | FALSE | FALSE | PRJEB30077 |
| 1036 | 1036_ToD_1_recurrent_pos | ToD | Spleen | recurrent | pos | 1 | 5+ | 2 | FALSE | FALSE | PRJEB30077 |
| 1037 | 1037_ToD_1_recurrent_pos | ToD | Spleen | recurrent | pos | 14 | 6+ | 2 | TRUE | FALSE | PRJEB30077 |
| 1037 | 1037_6-12m_2_recurrent_pos | 6-12m | Spleen | recurrent | pos | 15 | 6+ | 2 | TRUE | FALSE | PRJEB30077 |
| 1038 | 1038_ToD_1_primary_neg | ToD | Spleen | primary | neg | 0 | 4+ | 3 | FALSE | FALSE | PRJEB30077 |
| 1039 | 1039_ToD_1_primary_neg | ToD | Spleen | primary | neg | 0 | 5+ | 0.25 | FALSE | FALSE | PRJEB30077 |
| 1040 | 1040_ToD_1_recurrent_pos | ToD | Spleen | recurrent | pos | 10 | 6+ | 1 | TRUE | FALSE | PRJEB30077 |
| 1040 | 1040_EoT_2_recurrent_pos | EoT | Spleen | recurrent | pos | 11 | 3+ | 1 | TRUE | FALSE | PRJEB30077 |

| | | | | | | | | | | | |
|---|---|---|---|---|---|---|---|---|---|---|---|
| 1041 | 1041_ToD_1_primary_neg | ToD | BM | primary | neg | 0 | 2+ | 2 | FALSE | FALSE | PRJEB30077 |
| 1042 | 1042_ToD_1_primary_neg | ToD | Spleen | primary | neg | 0 | 2+ | 0.75 | FALSE | FALSE | PRJEB30077 |
| 1043 | 1043_ToD_1_primary_neg | ToD | Spleen | primary | neg | 0 | 5+ | 2 | FALSE | FALSE | PRJEB30077 |
| 1044 | 1044_ToD_1_primary_neg | ToD | Spleen | primary | neg | 0 | 2+ | 2 | FALSE | FALSE | PRJEB30077 |
| 1045 | 1045_ToD_1_recurrent_pos | ToD | Spleen | recurrent | pos | 3 | 6+ | 2 | TRUE | FALSE | PRJEB30077 |
| 1045 | 1045_6-12m_2_recurrent_pos | 6-12m | Spleen | recurrent | pos | 4 | 6+ | 2 | TRUE | FALSE | PRJEB30077 |
| 1045 | 1045_EoT-1_3_recurrent_pos | EoT-1 | Spleen | recurrent | pos | 4 | 4+ | 2 | TRUE | FALSE | PRJEB30077 |
| 1045 | 1045_EoT-2_4_recurrent_pos | EoT-2 | Spleen | recurrent | pos | 4 | 4+ | 2 | FALSE | TRUE | PRJEB30077 |
| 1046 | 1046_ToD_1_primary_neg | ToD | BM | primary | neg | 0 | 1+ | 1 | FALSE | FALSE | PRJEB30077 |
| 1047 | 1047_ToD_1_primary_neg | ToD | Spleen | primary | neg | 0 | -ve | 0.5 | FALSE | FALSE | PRJEB30077 |
| 1048 | 1048_ToD_1_primary_neg | ToD | Spleen | primary | neg | 0 | 4+ | 1 | FALSE | FALSE | PRJEB30077 |
| 1049 | 1049_ToD_1_primary_neg | ToD | BM | primary | neg | 0 | 3+ | 3 | FALSE | FALSE | PRJEB30077 |
| 1050 | 1050_ToD_1_primary_neg | ToD | BM | primary | neg | 0 | 2+ | 2 | FALSE | FALSE | PRJEB30077 |
| 1051 | 1051_ToD_1_primary_neg | ToD | Spleen | primary | neg | 0 | 5+ | 0.5 | FALSE | FALSE | PRJEB30077 |
| 1052 | 1052_ToD_1_primary_neg | ToD | Spleen | primary | neg | 0 | 3+ | 3 | FALSE | FALSE | PRJEB30077 |
| 1053 | 1053_ToD_1_primary_neg | ToD | BM | primary | neg | 0 | -ve | 1 | FALSE | FALSE | PRJEB30077 |
| 1054 | 1054_ToD_1_primary_neg | ToD | Spleen | primary | neg | 0 | 1+ | 0.5 | FALSE | FALSE | PRJEB30077 |

| | | | | | | | | | | | |
|---|---|---|---|---|---|---|---|---|---|---|---|
| 1055 | 1055_ToD_1_primary_neg | ToD | Spleen | primary | neg | 0 | 3+ | 1 | FALSE | FALSE | PRJEB30077 |
| 1056 | 1056_ToD_1_recurrent_pos | ToD | Spleen | recurrent | pos | 2 | 5+ | 1 | FALSE | FALSE | PRJEB30077 |
| 1057 | 1057_ToD_1_primary_neg | ToD | Spleen | primary | neg | 0 | 4+ | 2 | FALSE | FALSE | PRJEB30077 |
| 1058 | 1058_ToD_1_primary_neg | ToD | Spleen | primary | neg | 0 | 4+ | 0.5 | FALSE | FALSE | PRJEB30077 |
| 1059 | 1059_ToD_1_primary_neg | ToD | BM | primary | neg | 0 | -ve | 1 | FALSE | FALSE | PRJEB30077 |
| 1060 | 1060_ToD_1_primary_neg | ToD | BM | primary | neg | 0 | -ve | 2 | FALSE | FALSE | PRJEB30077 |
| 1061 | 1061_ToD_1_primary_neg | ToD | BM | primary | neg | 0 | 2+ | 0.75 | FALSE | FALSE | PRJEB30077 |
| 1062 | 1062_ToD_1_recurrent_pos | ToD | Spleen | recurrent | pos | 4 | 6+ | 1 | FALSE | FALSE | PRJEB30077 |
| 1062 | 1062_ToD_2_recurrent_pos | ToD | Spleen | recurrent | pos | 4 | 6+ | 1 | FALSE | TRUE | PRJEB30077 |
| 1063 | 1063_ToD_1_primary_neg | ToD | Spleen | primary | neg | 0 | 1+ | 3 | FALSE | FALSE | PRJEB30077 |
| 1064 | 1064_ToD_1_primary_neg | ToD | Spleen | primary | neg | 0 | 3+ | 1 | FALSE | FALSE | PRJEB30077 |
| 1065 | 1065_ToD_1_primary_neg | ToD | Spleen | primary | neg | 0 | 2+ | 0.5 | FALSE | FALSE | PRJEB30077 |
| 1066 | 1066_ToD_1_recurrent_pos | ToD | Spleen | recurrent | pos | 1 | 2+ | 1 | FALSE | FALSE | PRJEB30077 |
| 1067 | 1067_ToD_1_recurrent_pos | ToD | Spleen | recurrent | pos | 1 | 6+ | 2 | FALSE | FALSE | PRJEB30077 |
| 1068 | 1068_ToD_1_primary_neg | ToD | Spleen | primary | neg | 0 | 5+ | 1 | FALSE | FALSE | PRJEB30077 |
| 1069 | 1069_ToD_1_primary_neg | ToD | Spleen | primary | neg | 0 | 4+ | 2 | FALSE | FALSE | PRJEB30077 |
| 1069 | 1069_ToD_2_primary_neg | ToD | Spleen | primary | neg | 0 | 4+ | 2 | FALSE | TRUE | PRJEB30077 |

| | | | | | | | | | | | |
|---|---|---|---|---|---|---|---|---|---|---|---|
| 1070 | 1070_ToD_1_recurrent_pos | ToD | Spleen | recurrent | pos | 2 | 6+ | 0.5 | FALSE | FALSE | PRJEB30077 |
| 1071 | 1071_ToD_1_primary_neg | ToD | Spleen | primary | neg | 0 | -ve | 2 | FALSE | FALSE | PRJEB30077 |
| 1072 | 1072_ToD_1_primary_neg | ToD | Spleen | primary | neg | 0 | 3+ | 1 | FALSE | FALSE | PRJEB30077 |
| 1073 | 1073_ToD_1_primary_neg | ToD | Spleen | primary | neg | 0 | 1+ | 2 | FALSE | FALSE | PRJEB30077 |
| 1074 | 1074_ToD_1_primary_neg | ToD | Spleen | primary | neg | 0 | 5+ | 3 | FALSE | FALSE | PRJEB30077 |
| 1075 | 1075_ToD_1_recurrent_pos | ToD | Spleen | recurrent | pos | 3 | 6+ | 1 | FALSE | FALSE | PRJEB30077 |
| 1076 | 1076_ToD_1_primary_neg | ToD | Spleen | primary | neg | 0 | 4+ | 1 | FALSE | FALSE | PRJEB30077 |
| 1077 | 1077_ToD_1_recurrent_pos | ToD | Spleen | recurrent | pos | 5 | 6+ | 1 | FALSE | FALSE | PRJEB30077 |
| 1078 | 1078_ToD_1_recurrent_pos | ToD | Spleen | recurrent | pos | 11 | 5+ | 0.5 | TRUE | FALSE | PRJEB30077 |
| 1078 | 1078_6-12m_2_recurrent_pos | 6-12m | Spleen | recurrent | pos | 12 | 6+ | 0.5 | TRUE | FALSE | PRJEB30077 |
| 1079 | 1079_ToD_1_recurrent_pos | ToD | Spleen | recurrent | pos | 10 | 6+ | 2 | FALSE | FALSE | PRJEB30077 |
| 1079 | 1079_ToD_2_recurrent_pos | ToD | Spleen | recurrent | pos | 10 | 6+ | 2 | FALSE | TRUE | PRJEB30077 |
| 1080 | 1080_ToD_1_recurrent_pos | ToD | Spleen | recurrent | pos | 2 | 5+ | 2 | FALSE | FALSE | PRJEB30077 |
| 1080 | 1080_ToD_2_recurrent_pos | ToD | Spleen | recurrent | pos | 2 | 5+ | 2 | FALSE | TRUE | PRJEB30077 |
| 1081 | 1081_ToD_1_NA_neg | ToD | Spleen | NA | neg | NA | 2+ | 2 | FALSE | FALSE | PRJEB30077 |
| 1082 | 1082_ToD_1_recurrent_pos | ToD | Spleen | recurrent | pos | 14 | 6+ | 2 | FALSE | FALSE | PRJEB30077 |
| 1083 | 1083_ToD_1_recurrent_pos | ToD | Spleen | recurrent | pos | 1 | 6+ | 2 | FALSE | FALSE | PRJEB30077 |

| | | | | | | | | | | | |
|---|---|---|---|---|---|---|---|---|---|---|---|
| 1085 | 1085_ToD_1_primary_neg | ToD | BM | primary | neg | 0 | 3+ | 1 | FALSE | FALSE | PRJEB30077 |
| 1087 | 1087_NA_1_NA_NA | NA | NA | NA | NA | NA | NA | NA | FALSE | FALSE | PRJEB30077 |
| 1088 | 1088_ToD_1_primary_neg | ToD | Spleen | primary | neg | 0 | 1+ | 2 | FALSE | FALSE | PRJEB30077 |
| 1089 | 1089_ToD_1_primary_neg | ToD | Spleen | primary | neg | 0 | 2+ | 2 | FALSE | FALSE | PRJEB30077 |
| 1091 | 1091_ToD_1_primary_neg | ToD | Spleen | primary | neg | 0 | 3+ | 1 | FALSE | FALSE | PRJEB30077 |
| 1092 | 1092_ToD_1_primary_neg | ToD | Spleen | primary | neg | 0 | 2+ | 1 | FALSE | FALSE | PRJEB30077 |
| 1093 | 1093_ToD_1_primary_neg | ToD | Spleen | primary | neg | 0 | 3+ | 1 | FALSE | FALSE | PRJEB30077 |
| 1094 | 1094_ToD_1_primary_neg | ToD | BM | primary | neg | 0 | 2+ | 0.5 | FALSE | FALSE | PRJEB30077 |
| 1095 | 1095_EoT_1_primary_neg | EoT | Spleen | primary | neg | 0 | 3+ | 1 | FALSE | FALSE | PRJEB30077 |
| 1096 | 1096_ToD_1_primary_pos | ToD | Spleen | primary | pos | 0 | 4+ | 1 | FALSE | FALSE | PRJEB30077 |
| 1097 | 1097_6-12m_1_recurrent_pos | 6-12m | Spleen | recurrent | pos | 4 | 6+ | 6 | FALSE | FALSE | PRJEB30077 |
| 1098 | 1098_ToD_1_recurrent_pos | ToD | Spleen | recurrent | pos | 2 | 6+ | 0.75 | FALSE | FALSE | PRJEB30077 |
| 1099 | 1099_ToD_1_recurrent_pos | ToD | Spleen | recurrent | pos | 3 | 1+ | 3 | FALSE | FALSE | PRJEB30077 |
| 1100 | 1100_ToD_1_primary_pos | ToD | Spleen | primary | pos | 0 | 6+ | 0.5 | FALSE | FALSE | PRJEB30077 |
| 1101 | 1101_6-12m_1_recurrent_pos | 6-12m | Spleen | recurrent | pos | 2 | 6+ | 0.5 | FALSE | FALSE | PRJEB30077 |

**Table S2.** Aneuploidy profiles and metadata summary of all isolates from patients with time series data and /or replicates of primary isolates.

| Profile identifier | Profile count | Patient code | Sample [patient code, study timepoint, sample no for patient, VL category, HIV status] | Aneuploidy profile [differences to most common profile: all chrs are 2, except chr 31 with 4] | Replicate of primary isolate |
|---|---|---|---|---|---|
| 1 | 83 | 1004 | 1004_ToD_1_recurrent_pos | ------------------------------------ | FALSE |
| 1 | 83 | 1004 | 1004_6-12m_2_recurrent_pos | ------------------------------------ | FALSE |
| 16 | 1 | 1023 | 1023_EoT_1_primary_pos | --1------1-------------------------- | FALSE |
| 2 | 9 | 1023 | 1023_3m_2_recurrent_pos | ------------------------------1----- | FALSE |
| 2 | 9 | 1023 | 1023_6-12m_3_recurrent_pos | ------------------------------1----- | FALSE |
| 2 | 9 | 1023 | 1023_EoT_4_recurrent_pos | ------------------------------1----- | FALSE |
| 2 | 9 | 1026 | 1026_ToD_1_recurrent_pos | ------------------------------1----- | FALSE |
| 2 | 9 | 1026 | 1026_6-12m_2_recurrent_pos | ------------------------------1----- | FALSE |
| 1 | 83 | 1037 | 1037_ToD_1_recurrent_pos | ------------------------------------ | FALSE |
| 2 | 9 | 1037 | 1037_6-12m_2_recurrent_pos | ------------------------------1----- | FALSE |
| 2 | 9 | 1040 | 1040_ToD_1_recurrent_pos | ------------------------------1----- | FALSE |
| 2 | 9 | 1040 | 1040_EoT_2_recurrent_pos | ------------------------------1----- | FALSE |
| 1 | 83 | 1045 | 1045_ToD_1_recurrent_pos | ------------------------------------ | FALSE |
| 12 | 1 | 1045 | 1045_6-12m_2_recurrent_pos* | ------1---------------1-------1----- | FALSE |
| 14 | 1 | 1045 | 1045_EoT-1_3_recurrent_pos* | ------1--1------------1-------1----- | FALSE |
| 13 | 1 | 1045 | 1045_EoT-2_4_recurrent_pos* | ------1--1---------------11---1----- | TRUE |
| 5 | 2 | 1062 | 1062_ToD_1_recurrent_pos | --------------------------1-----1--- | FALSE |
| 5 | 2 | 1062 | 1062_ToD_2_recurrent_pos | --------------------------1-----1--- | TRUE |
| 1 | 83 | 1069 | 1069_ToD_1_primary_neg | ------------------------------------ | FALSE |
| 1 | 83 | 1069 | 1069_ToD_2_primary_neg | ------------------------------------ | TRUE |
| 3 | 5 | 1078 | 1078_ToD_1_recurrent_pos | ------1----------------------------- | FALSE |
| 3 | 5 | 1078 | 1078_6-12m_2_recurrent_pos | ------1----------------------------- | FALSE |
| 1 | 83 | 1079 | 1079_ToD_1_recurrent_pos | ------------------------------------ | FALSE |
| 1 | 83 | 1079 | 1079_ToD_2_recurrent_pos | ------------------------------------ | TRUE |
| 1 | 83 | 1080 | 1080_ToD_1_recurrent_pos | ------------------------------------ | FALSE |
| 1 | 83 | 1080 | 1080_ToD_2_recurrent_pos | ------------------------------------ | TRUE |
| * reinfection compared to first sample of this patient | | | | | |
| Negative differences to the dominant aneuploidy profile are circled in red. | | | | | |
| Row in gray indicate strain data with aneuploidy between timeseries or aliquot sampling. | | | | | |
